# Exhaustive circulating tumor DNA sequencing reveals the genomic landscape of Hodgkin lymphoma and facilitates ultrasensitive detection of minimal residual disease

**DOI:** 10.1101/2021.03.16.21253679

**Authors:** Sophia Sobesky, Laman Mammadova, Melita Cirillo, Esther Drees, Julia Mattlener, Helge Dörr, Janine Altmüller, Zhiyuan Shi, Paul J Bröckelmann, Jonathan Weiss, Stefanie Kreissl, Stephanie Sasse, Roland T Ullrich, Sarah Reinke, Wolfram Klapper, Elena Gerhard-Hartmann, Andreas Rosenwald, Margaretha GM Roemer, Peter Nürnberg, Anton Hagenbeek, Josée M. Zijlstra, Dirk Michiel Pegtel, Andreas Engert, Peter Borchmann, Bastian von Tresckow, Sven Borchmann

## Abstract

Individualizing treatment is key to improve outcome and reduce long-term side-effects in any cancer. In Hodgkin lymphoma (HL), individualization of treatment is hindered by a lack of genomic characterization and technology for sensitive, molecular response assessment.

Sequencing of cell-free (cf)DNA is a powerful strategy to understand an individual cancer genome and can be used to develop assays for extremely sensitive disease monitoring. In HL, a high proportion of cfDNA is tumor-derived making it a highly relevant disease model to study the role of cfDNA sequencing in cancer.

Here, we introduce our targeted cfDNA sequencing platform and present the largest genomic landscape of HL to date, which was entirely derived by cfDNA sequencing. We comprehensively genotype and assess minimal residual disease in 324 samples from 121 patients, presenting an integrated landscape of mutations and copy number variations in HL. In addition, we perform a deep analysis of mutational processes driving HL, investigate the clonal structure of HL and link several genotypes to HL phenotypes and outcome. Finally, we show that minimal residual disease assessment by repeat cfDNA sequencing as early as a week after treatment initiation is feasible and predicts overall treatment response allowing highly improved treatment guidance and relapse prediction. Our study also serves as a blueprint showcasing the utility of our platform for other cancers with similar therapeutic challenges.

## Introduction

Classical Hodgkin lymphoma (HL) is a B-cell derived malignant lymphoma that is now often curable with aggressive chemotherapy^1, 2^. Nevertheless, many challenges exist. Aggressive, multi-agent front-line chemo- and radiotherapy leads to early- and late-toxicities such as secondary cancers, cardiovascular disease, infertility, fatigue and osteonecrosis^3–7^. Furthermore, 10-30% of patients fail initial treatment and many of these often young patients ultimately die^8^.

Individualizing treatment is key to improve outcome and reduce long-term side-effects in any cancer. Principally, two strategies for treatment individualization in a curable cancer exist: upfront treatment individualization by accurate risk assessment or accurate assessment of treatment response to stop treatment as soon, as cure is achieved.

In HL, both strategies are currently hampered by a lack of genomic characterization of the disease and sensitive, molecular response assessment. Upfront risk assessment is currently purely based on clinical risk factors such as disease extent and has not improved much in recent decades^9^. Biological risk classification is impeded, because a comprehensive genomic characterization of HL has so far been challenging due to the paucity of malignant Hodgkin Reed-Sternberg (HRS) cells in the typical HL tumor biopsy^10^. Past studies relied on cumbersome and technically challenging laser microdissection or flow sorting of biopsies for enrichment of HL cells^11–14^. Response assessment in HL is currently based on positron emission tomography (PET). However, sensitivity and specificity of PET-based response assessment is limited^15–17^.

In contrast, biological risk classifications based on the detection of genetically defined subgroups has superseded clinical risk classification systems and improved outcome in other cancers such as acute myeloid leukemia^18^. Likewise, highly sensitive, molecular response assessment based on the detection of minimal residual disease (MRD) has greatly improved outcome in acute lymphoblastic leukemia^19^.

A powerful strategy to unravel the cancer genome of a patient is the sequencing of cell-free DNA (cfDNA), which is released into the circulation from apoptotic cells and contains circulating tumor-derived DNA (ctDNA) with mutations, copy number alterations, gene fusions or specific (clonal) immune-receptor (IgH) rearrangements representative of the tumor^20, 21^. We and others have found that the plasma ctDNA content is much higher than one would expect in HL^21–24^, considering the paucity of tumor cells, making it a highly relevant disease model to study ctDNA. Specifically, sequencing of ctDNA in HL allows easy and clinically feasible genotyping and biological risk assessment. Furthermore, carefully developed strategies to reduce sequencing errors enable highly sensitive and specific minimal residual disease assessment by sequencing of ctDNA^25, 26^.

Here, we comprehensively genotype and assess minimal residual disease in 324 longitudinally collected samples from 121 HL patients before, during and after treatment. We present the largest integrated landscape analysis of mutations and copy number variations in HL to date and perform deep analysis of mutational processes driving HL, associations between HL and viruses and bacteria and clonal structure of HL. We link several genotypes to HL phenotypes and treatment outcome and show that MRD assessment is informative for overall treatment response as early as one week after treatment initiation suggesting high clinical utility.

## Results

### Technical and clinical validation

We aimed at designing a targeted ctDNA sequencing and bioinformatics platform for accurate baseline genotyping and MRD detection during follow-up. To this end, cfDNA sequencing artifacts and stochastic sequencing errors need to be suppressed^25^. Combining existing knowledge^22, 23, 25^, our error suppression approach includes tagging DNA molecules with unique molecular identifiers (UMIs) and a combination of two comparative error suppression (CES) methods: (I) deriving and excluding error-prone genomic regions in a identically processed set of cfDNA samples from a healthy control cohort and (II) judging each base call based on its modelled error-rate in an identically processed healthy control cohort considering the given tri-nucleotide context^27^ (see methods for details).

For technical validation we used spike-ins of well-characterized reference DNA (NA12878)^28, 29^ fragmented to cfDNA-size into healthy donor cfDNA. Observed error profiles depended on use of UMI-based error suppression or additional CES (Fig. 1A-C). Combined UMI-based and CES error suppression resulted in improved sensitivity and specificity of mutation calling at 0.5% spiked-in mutated allele frequency (mAF) (Fig. 1D-E). UMIs contributed most to sensitivity (Fig. 1D) whereas CES was essential for high specificity (Fig. 1E). For technical validation of MRD detection we used lower spike-ins of 0.5%, 0.05%, 0.025%, 0.0125% and 0.005% mAF and were able to show that combined error-suppression is necessary to reliably distinguish MRD from background below a mAF (corresponding to the MRD level) of 0.025% or 1 mutated allele in 4000 DNA molecules (Fig. 1F).

**Figure 1:**
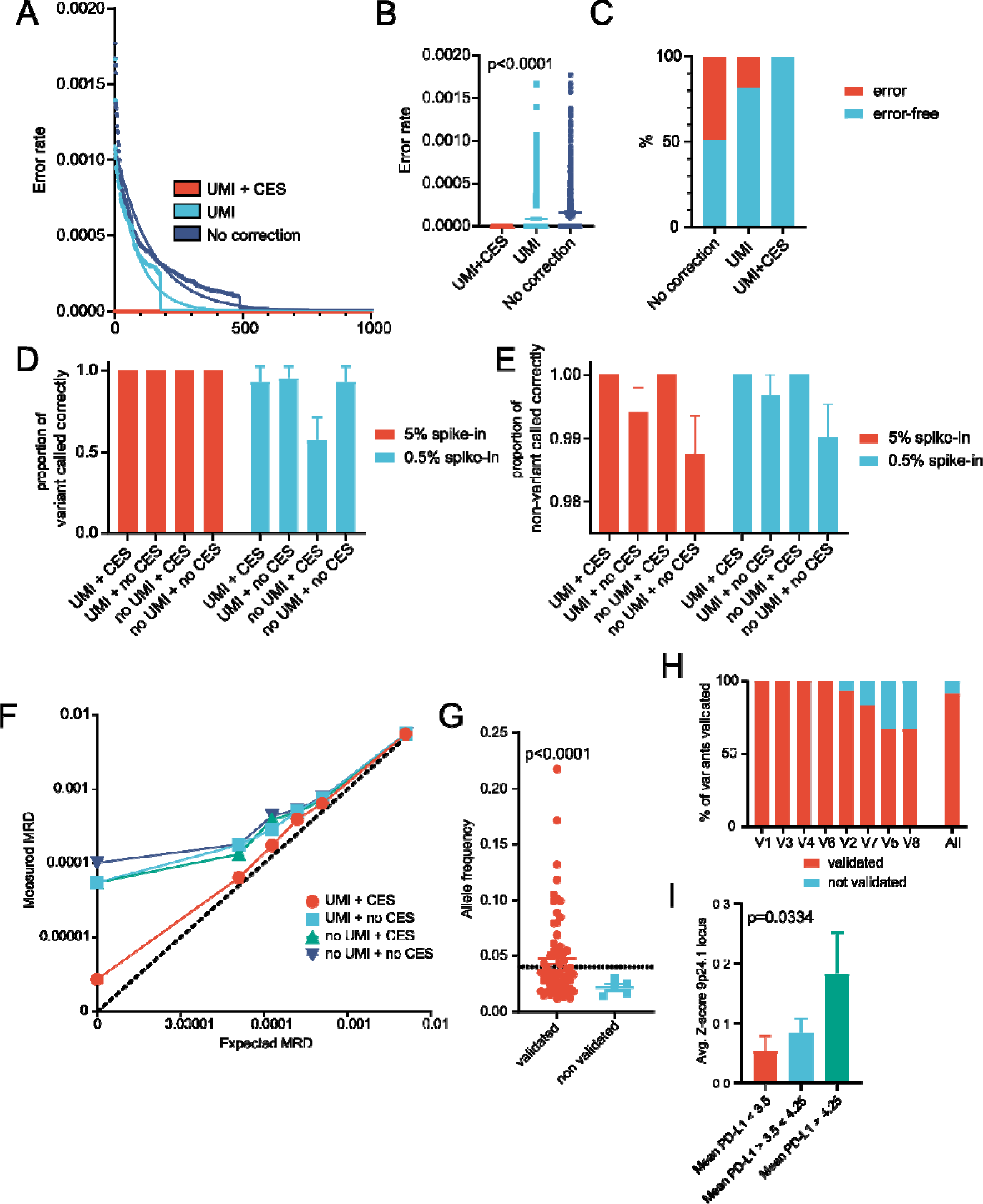
Technical and clinical validation of ctDNA sequencing platform. (A) error profile of indicated error suppression methods across 1000 random non-mutated bases. Dots indicate observed error rate at a base. Lines indicate a one-phase decay model of the respective error rate. (B) average error rate of indicated error suppression methods across 1000 random non-mutated bases, (C) error-free rate of indicated error suppression methods across 1000 random non-mutated bases, (D-E) technical validation of variant calling: shown is the proportion of correctly called mutations across 42 truly mutated bases (D) and the proportion of correctly called non-mutated bases across 1527 random non-mutated bases (E) for the indicated error suppression methods, (F) technical validation of minimal residual disease assessment with linearity test for indicated error suppression methods (n=24). The dotted line indicates true linearity (the measured allele frequency (AF) in technical spike-in controls equals the expected (spiked-in) AF (G) comparison between AF of variants identified in cfDNA that were validated in FFPE-tissue and those that were not (n=8 clinical validation samples), (H) validation rate of variants identified in cfDNA in FFPE tissue samples shown separately across 8 sample pairs and summarized for all sample pairs, (I) comparison between HRS cell CD274/PD-L1 copies assessed by FISH in tissue and copy number z-score in corresponding cfDNA samples (n=51). Error bars show bootstrap confidence intervals for (D) and (E) and s.e.m. for (B), (G) and (I). One-way ANOVA (B), t-test with Welch correction (G) and one-way ANOVA with trend test (I) were used.

For clinical validation of mutation calling, we compared detected variants in 8 cfDNA samples with corresponding Formalin-Fixed Paraffin-Embedded (FFPE) tumor biopsies. We confirmed all mutations called in cfDNA with a mAF > 4% (26/26) and 91.3% (63/69) of all mutations irrespective of mAF (Fig. 1G-H). This is in line with expectations, as mutation calling in artifact rich bulk FFPE samples with low HL cell content will fail to detect rare variants, underscoring the value of ctDNA sequencing in HL. For clinical validation of copy number assessment in cfDNA, we compared 9p24.1 (locus of PD-L1) gains assessed in cfDNA with gold standard Fluorescence in situ hybridization (FISH). Average PD-L1 copies assessed by FISH in HL lymph node biopsies correlated strongly with copy number gain in cfDNA (Fig. 1I). Based on standard protocols for assay validation^30^ we defined a limit of blank of 0.00714% (appx. 1 in 14,000 DNA molecules) and a limit of detection of 0.00945% (appx. 1 in 10,500 DNA molecules) for MRD detection. Employing baseline variant calls from 6 HL patients, we tested 10 healthy donor samples as negative MRD controls. All healthy control samples showed no evidence of MRD (Supplementary Table 1).

### Sequencing and patient characteristics

We produced a mean of 2.02×10^8^ (±7.51×10^6^ standard error of mean) reads per HL patient cfDNA sample with a mean of 6.84×10^7^ (±3.65×10^6^) uniquely mapped reads (Fig. S1A). The high duplication rate (mean: 3.67±0.25) (Fig. S1B) was intentional and utilized for UMI-based error suppression. Mean post UMI-deduplication per gene coverage was 1902x (±48) and mean per patient coverage was 1670x (±90) (Fig. S1C-D). The median cfDNA concentration was 14.1 ng/ml plasma (range: 2.5-557.5 ng/ml) (Fig. S1E). Baseline cfDNA concentration was higher in males (median 19.7 vs. 10.8 ng/ml, p=0.0587) (Fig. S1F) and patients with higher clinical stage (p=0.0001) (Fig. S1G) or higher International Prognostic Score^9^ (p<0.0001) (Fig. S1H). Patient characteristics were as expected with a representative stage, age, gender and histological subtype distribution^1^ (Supplementary Table 2).

### Mutational landscape of Hodgkin lymphoma

We detected 4738 single base substitutions (SBS) and small InDels in our cohort with a median of 36 mutations per patient (range 1-175) (Fig. S1I, Supplementary Table 3). Variants were identified in 109 out of 111 patients (98%). Nonsynonymous mutations were predominant (n=2050), followed by non-coding mutations (n=1891), synonymous mutations (n=614), mutations in untranslated regions (UTRs) (n=92) and splicing mutations (n=44) (Fig. S1J). This was as expected for our enrichment approach targeting mostly exons.

The genes with most non-synonymous, non-intronic and non-intergenic mutations were TTN (n=71), SOCS1 (n=58), TNFAIP3 (n=51), ITPKB (n=50), STAT6 (n=42), GNA13 (n=36), B2M (n=34) and CSF2RB (n=28) (Supplementary Table 4) (Fig. S2A-B), after conservatively removing putative mutations detected in MUC4 or MUC16 as these could be artifacts resulting from paralogous alignment^31^. The number of mutations per patient was highly heterogeneous (Fig. S2C-D).

Mutational frequency is not an indicator of relevance, for example because large genes such as TTN, the most frequently mutated gene in this cohort, possibly only harbors many mutations because of its size^31^. To identify genes likely relevant in HL pathogenesis and not mere bystander mutations we used a modified Vogelstein rule^32^ (see methods for details). We identified 23 tumor suppressor genes (TSG) and 20 oncogenes (OG) (Fig. 2A, Supplementary Table 5). The number of OG and TSG mutations per patient was highly heterogeneous with a median of 5 (Fig. 2B-C). We identified recurrent mutations in several known oncogenic pathways in HL such as JAK-STAT signaling^33^ (SOCS1, STAT6, CSF2RB, IL4R), Class I MHC mediated antigen processing and presentation^12^ (B2M) and NF-kappa B signaling^34^ (TNFAIP3, IKBKB, NFKBIE). In addition, we identified several recurrent TSGs and OGs involved in chromatin modification (EZH2, ARID1A, PBRM1), ubiquitination (UBE2A, FBXO11), cAMP signaling (GRIN2A, PLCE1, GNAI2), transcriptional cell fate regulation by TP53 (TNRC6B, FAS, CDKN2A), cell cycle regulation (XPO1, CCND3) and transcriptional misregulation in cancer (JMJD1C, MYCN, TCF3), interferon signaling (IRF4, IRF8, PTPN1) as well as histone genes (HIST1H1E, HIST1H2AM) (Fig. 2D). Next, we expanded our analysis by allocating identified non-synonymous mutations to these pathways based on predefined gene sets (Supplementary Table 6). Almost all patients had histone mutations (87.9%), while most patients had mutations in the JAK-STAT signaling pathway (71.0%), interferon signaling (60.7%), ubiquitination (60.7%), Class I MHC mediated antigen processing and presentation (57.9%) and NF-kappa B signaling (52.3%) (Fig.2D).

**Figure 2:**
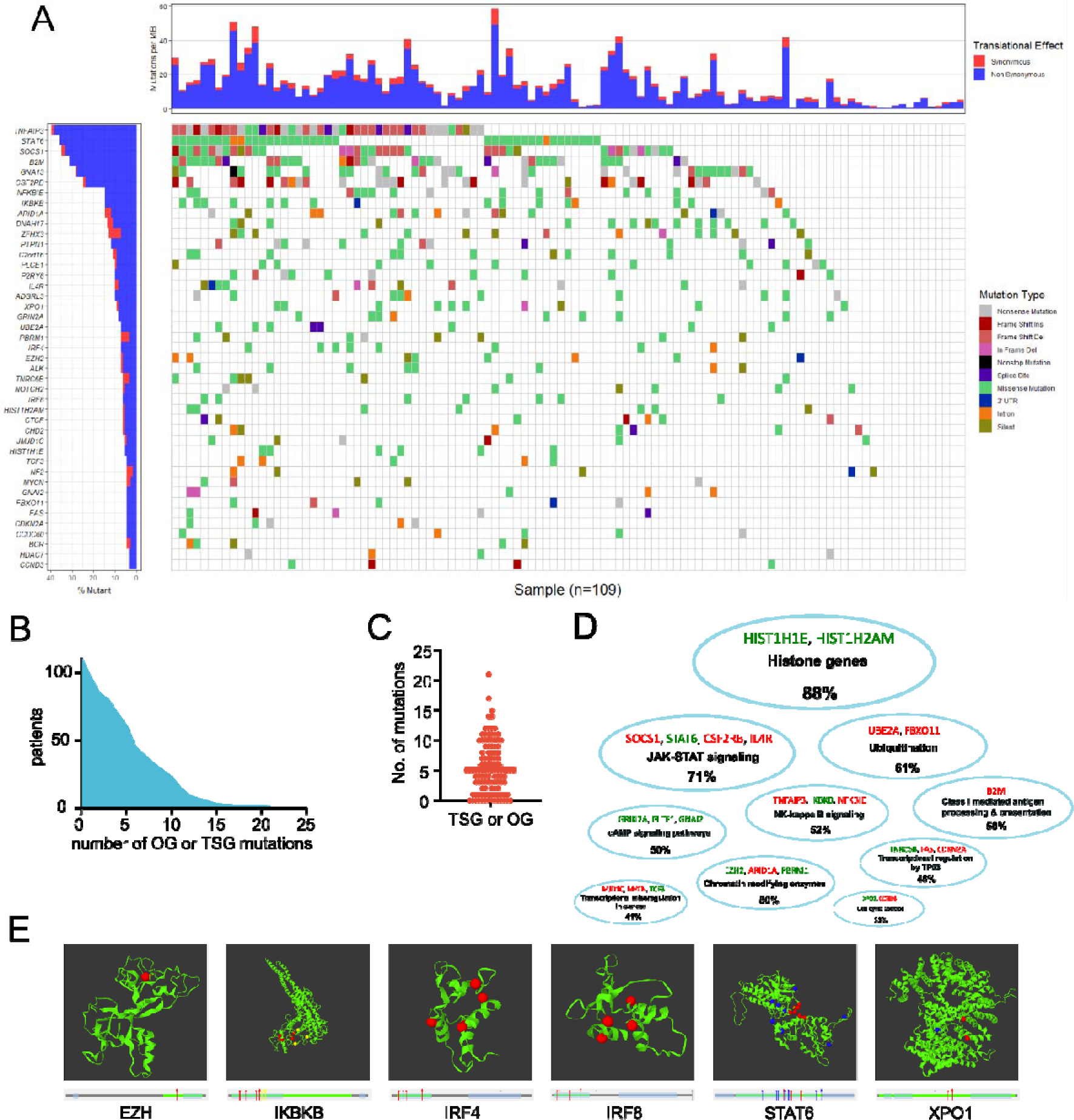
Mutational landscape of Hodgkin lymphoma. (A) waterfall plot of oncogenes and tumor suppressor genes identified in the patient cohort (n=109, 2 patients without identified mutations not shown). The chart on the left shows the genes and proportions of patients with mutations. The chart on top shows the mutational burden per patient. Color codes indicate the type of mutation in a patient and proportion of synonymous vs. non-synonymous mutations in genes (left chart) and patients (top chart), respectively. (B) cumulative plot of patients and number of mutations in identified oncogenes and tumor suppressor genes (n=111), (C) number of mutations in identified oncogenes and tumor suppressor genes per patient (n=111), (D) cloud figure of affected pathways by mutations in oncogenes and tumor suppressor genes. Percentages indicate proportion of patients affected by at least one mutation in indicated pathway. Shown identified oncogenes (green) and tumor suppressor genes (red) are color coded, (E) 3D protein models of oncogenes with significant spatial clustering of mutations. Red, blue, and yellow dots indicate separate spatial clusters. The charts below the 3D models depict linear representations in which arrows with the same colors as in the 3D models indicate position of spatial clusters on this linear representation of the protein sequence. Error bars show s.e.m.

To understand their functional relevance, we analyzed spatial clustering of mutations in OGs within 3-dimensional protein structures using Mutation3D^35^. We identified significant spatial clustering in EZH2, IKBKB, IRF4, IRF8, STAT6 and XPO1 (Fig. 2E). Mutations in EZH2 all occurred in residue 641 (NM_001203247) (p_cluster_=6.67×10^-5^). Recurrent Y641 EZH2 mutations in HL have so far not been reported^11^. For IKBKB, we identified two clusters (p_cluster_=1.7×10^-3^ each) in the PKinase domain of IKBKB with possible gain-of-function and increased NF-κB signaling^36^. For IRF4 we identified a cluster in the IRF (DNA binding) domain (p_cluster_=1.83×10^-4^). Recurrent IRF4 mutations have not yet been described in HL. Mutations in the identified cluster are in close proximity to mutations that have been shown to increase IRF4 transcriptional activity^37^. Similar to IRF4, we identified a cluster of mutations in IRF8 in the IRF (DNA binding) domain (p_cluster_=2.52×10^-4^). For STAT6 we identified a cluster of mutations in the STAT (DNA binding) domain (p_cluster_=5.56×10^-2^). Functionally, STAT6 expression is important for HL survival^38^. Finally, we identified a cluster of mutations in XPO1 (p_cluster_=9.71×10^-5^) consisting mostly of the recurrent E571K (NM_003400) mutation. Functionally, it has been shown that the XPO E571K mutation is a gain-of-function mutation leading to clonal B-cell malignancy^39^. A more detailed functional discussion of the identified mutation clusters is available as a supplementary note.

Furthermore, we identified several co-occurring or mutually exclusive OGs and TSGs using DISCOVER^40^ (Fig. S3A-B). However, co-occurrence or mutual exclusivity analysis did not reveal any clear relationships between pathways beyond the single gene relationships outlined in Fig. S3.

### Mutational signatures

Mutational signatures have been identified as characteristic processes of somatic mutation acquisition in cancer reflecting distinct mechanisms^41^. We identified 6 distinct COSMIC single base substitution (SBS) signatures^41^ (Fig. 3A) using deconstructSigs^42^: SBS1 (deamination of 5-methylcytosine to thymine, associated with aging), SBS3 (defective homologous recombination-based (HR) DNA damage repair), SBS6 and SBS15 (defective DNA mismatch repair (MMR), associated with microsatellite instability) SBS9 (somatic hypermutation in lymphoid cells) and SBS25 (unknown cause, hitherto only identified in HL cell lines)^43^ (Fig. 3B).

**Figure 3:**
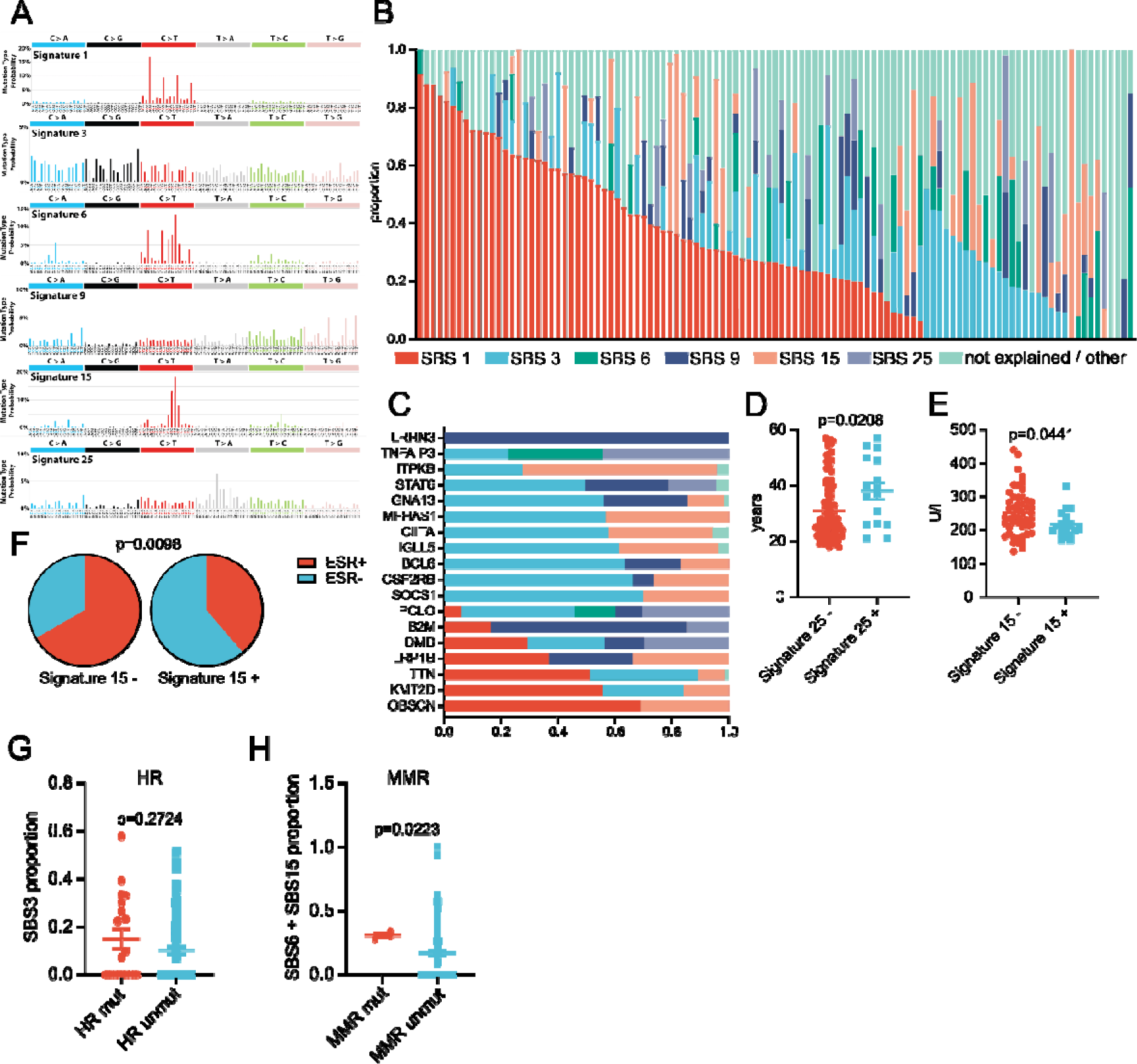
Mutational signatures in Hodgkin lymphoma. (A) mutational profile of mutational signatures identified in Hodgkin lymphoma (Figure adapted from https://cancer.sanger.ac.uk/cosmic/signatures/SBS/). (B) contribution of identified mutational signatures to single patients sorted by contribution of SBS1 (n=109), (B) contribution of identified mutational signatures to indicated genes, (D) comparison of patient age between patients with or without significant detection of SBS25 (n=109), (E) comparison of lactate dehydrogenase (LDH) at diagnosis between patients with or without significant detection of SBS 5 (n=68), (F) comparison of elevated erythrocyte sedimentation rate (ESR) between patients with or without significant detection of SBS15 (n=109), (G) comparison of SBS3 contribution between patients with and without mutations in genes involved in homologous recombination, (H) comparison of combined SBS6 + SBS15 contribution between patients with and without mutations in genes involved in mismatch repair. Error bars show s.e.m. T-test (D-E), Fisher’s exact test (F) and Mann-Whitney test (G-H) were used.

Next, we examined the contribution of these mutational signatures to mutations in recurrently mutated genes in our cohort. We identified distinct mutational processes active in different genes. SBS1 was the biggest contributor in DMD, LRP1B, TTN, KMT2D and OBSCN; SBS3 the biggest contributor in STAT6, GNA13, MFHAS1, CIITA, IGLL4, BCL6, CSF2RB and SOCS1; SBS15 the biggest contributor in ITPKB; SBS9 the biggest contributor in B2M and LRRN3; and SBS25 the biggest contributor in TNFAIP3 (Fig. 3C). Overall, SBS1 - the aging signature - was the main contributor in genes that are not OGs or TSGs, suggesting a role of the underlying process in inducing bystander mutations. Interestingly, SBS3 - the defective homologous recombination signature - contributed majorly to mutations in genes crucial for HL oncogenesis, such as STAT6, CSF2RB or SOCS1^12, 33^, highlighting a potential role for defective homologous recombination in HL pathogenesis. SBS9 – the somatic hypermutation signature – was the sole contributor to LRRN3 mutations, which has not been empirically shown to be a target of somatic hypermutation but predicted to be one in one analysis^44^. SBS25 – a potential HL specific signature - was the main contributor to mutations in TNFAIP3, one of the most frequently mutated genes in HL^45^, supporting the notion that SBS25 is indeed a HL signature and not an artifact.

Correlating mutational signatures with patient phenotypes, we observed that patients with detection of SBS25 were older (38.1 vs. 31.0 years of age, p=0.0208) (Fig. 3D), while detection of SBS15 was linked to lower lactate dehydrogenase (LDH) levels (217 vs. 253 U/l, p=0.0441) (Fig. 3E) and less frequent erythrocyte sedimentation rate (ESR) elevation (OR=0.32, p=0.0098) (Fig. 3F), suggesting SBS15 could be associated with a less inflammatory and lower tumor burden or tumor cell turnover phenotype.

Next, we examined if mutations in genes involved in homologous recombination (HR) or mismatch repair (MMR) (Supplementary table 7) were associated with SBS3 (defective HR) or SBS6 and SBS15 (defective MMR), respectively. The contribution of SBS3 was higher in patients with mutations in genes involved in HR, although this was not significant (Fig. 3G). The combined contribution of SBS6 and SBS15 was significantly higher in patients with mutations in genes involved in MMR (Fig. 3H). These results suggest that subsets of HL exist with defects in HR, MMR or both and that the respective mutational process contributes fittingly to the mutations observed in these patient subsets.

### Somatic copy number variations

We used cnvkit^46^ and Gistic 2.0^47^ to identify focal and arm-level recurrent somatic copy number variations (sCNV). Overall, we identified 16 recurrent gains and 34 losses (q<0.25) (Fig. 4A-B, Fig. S4A). The most common recurrent gains were 19p13.2 (65%), 2q31.2 (62%), 12q13.3 (STAT2, STAT6) (62%), 2p16.1 (REL) (61%), and 9p24.1 (JAK2, CD274) (55%). We also observed frequent gains of regions including NFKB1, MLL5 and PIK3CG (Fig. 4C). The most common recurrent losses were 6q23.3 (TNFAIP3, ECT2L) (61%), 9q13 (59%), 13q32.3 (59%), 6q22.31 (58%) and 15q15.1 (57%). Of note, we also observed frequent losses of regions including B2M, HIST1H1E, HIST1HIB, and the HLA locus, underscoring the functional relevance of histone genes and immune escape in HL (Fig. 4D). A separate, arm-level analysis using Gistic 2.0^47^, was consistent with these findings, identifying 2p, 2q, 5p, 9p, 10p, 12p, 12q as recurrent arm-level gains and, among others, 4q, 6p, 6q, 7q, 11q, 17p, 18p, 18q and 22q as recurrent arm-level gains. Both, recurrent, arm-level gains and losses, were observed on 17q, 19p and 19q. (Fig. S4B).

**Figure 4:**
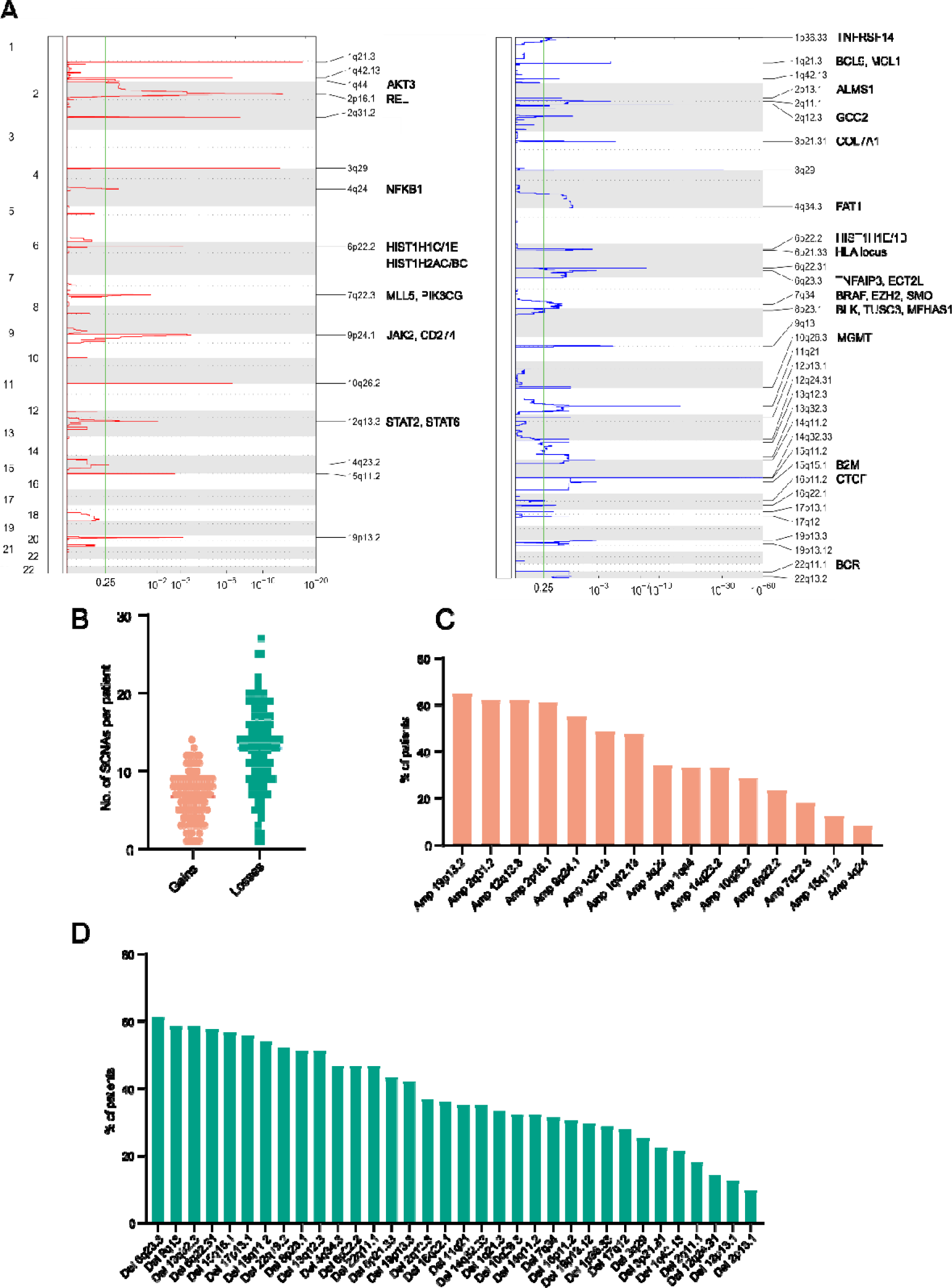
Copy number variations in Hodgkin lymphoma. (A) Recurrent copy number gains (red) and losses (blue) in patient cohort (n=111). The x-axis shows the chromosomes, the y-axis shows the GISTIC q-values. Each peak indicates a recurrent copy number gain or loss with the width of the peak corresponding to the size of the recurrent region and the height to the q-value. Selected genes within recurrent regions are annotated, (B) distribution of number of detected copy number gains and losses in patient cohort (n=111), (C) most frequently detected recurrent copy number gains in patient cohort (n=111), (D) most frequently detected copy number losses in patient cohort (n=111).

### Detection of viral and bacterial associations

We used a previously developed pipeline^48^ built around KRAKEN^49^ to identify viral and bacterial associations with HL by liquid biopsy. We detected Epstein-Barr-Virus (EBV) and Human Herpesvirus 6B (HHV6B), both known to be associated with HL^50^, in 17 (15.3%) and 7 (6.3%) patients, respectively (Fig. S5A-B). Validating our findings, we found that EBV DNA in plasma as measured by our pipeline corresponded very well to EBV status of the tumor cells assessed by LMP1 staining of infiltrated HL lymph nodes (Fig. S5C). HL cases associated with EBV were older (37.2 vs. 31.1 years of age, p=0.0414) (Fig. S5D) and less likely to have mutations in genes involved in NF-kappa B signaling (OR=0.35, p=0.0696) (Fig. S5E), in line with previous reports describing histological EBV detection associated with older HL patients^51^ and fewer mutations in TNFAIP3^34^. Also, our observation may be related to the notion that EBV-infected B-cells rely on NFkB for survival^52^. In contrast to previous reports^14, 33^, we detected no association between mutational burden and EBV in our cohort (6.0 vs. 7.3 mutations/Megabase (Mb), EBV+ vs. EBV-, p=0.3418).

HL cases associated with HHV6B were older (42.2 vs. 31.3 years of age, p=0.0144) (Fig. S5F), more likely to have extranodal involvement (OR=5.17, p=0.0443) (Fig. S5G) and had a higher proportion of SBS6 detection (OR=6.22, p=0.0105) (Fig. S5H). The identified associations of HHV6B with a distinct HL phenotype have not been described previously. Of note, EBV and HHV6B were not detected in any of the healthy control or matched germline samples (Supplementary Table 8).

We also detected a number of bacterial taxa in the plasma of HL patients after extensive filtering to exclude false positives (see methods for details): *Streptococcus mitis*, *Burkholderia cepacia*, *Dolosigranulum pigrum*, *Mycobacteroides salmoniphilum*, *Ruminococcus gnavus* and *Neisseria mucosa* (Supplementary Table 9, Fig. S5I). It is possible that detection of some species, such as *Streptococcus mitis* as a common colonizer of humans, was increased in cfDNA due to HL-mediated immunosuppression^53^. Of note we also detected *Moraxella catarrhalis* in a single plasma sample with no detection in the matched germline sample (Supplementary Table 10). An uncontrolled IgD+ B cell response to *Moraxella catarrhalis* infection has recently been described as a possible early oncogenic driver in nodular lymphocyte-predominant Hodgkin lymphoma^54^.

### HL phenotypes are characterized by distinct genotypes

Our cohort included 40 patients with a large mediastinal mass. These patients were characterized by a higher frequency of B2M (OR=4.39, p=0.0009) (Fig. S6A) and IKBKB (OR=3.78, p=0.0337) (Fig. S6B) mutations, as well as more mutations in genes involved in Interferon signaling (OR=3.05, p=0.0095) (Fig. S6C) and JAK-STAT signaling (OR=2.43, p=0.0577) (Fig. S6D). These alterations have a high overlap with those recently reported in primary mediastinal B-cell lymphoma (PMBL)^55^. Taken together with the notion that a disease continuum stretching from mediastinal HL via grey zone lymphoma to PMBL exists^56^, these findings are suggestive of a biological subgroup of HL with a proclivity to mediastinal growth biologically and phenotypically close to PMBL.

Probing age-genotype associations, we found EZH2 mutations to be associated with younger (21.8 vs. 32.4 years, p<0.0001) (Fig. S6E) and IRF4 mutations to be associated with older patients (44.8 vs. 31.3 years, p=0.0488) (Fig. S6F).

Furthermore, patients with any mutations in genes involved in transcriptional regulation by p53 (267 vs. 226 U/l, p=0.0111) (Fig. S6G) had higher baseline LDH levels, suggesting higher tumor burden or tumor cell turnover in these patients.

### Association between HL genotypes and treatment response

Biological predictors of response are currently not established in HL. Therefore, we examined associations of somatic mutations and sCNVs with early PET response after 2 treatment cycles. Mutations in IRF8 (OR=5.97, p=0.0324) (Fig. S7A), PIM1 (OR=5.97, p=0.0324) (Fig. S7B), TP53 (OR=4.88, p=0.0280) (Fig. S7C) and NOTCH1 (OR=4.63, p=0.0799) (Fig. S7D) were associated with inferior response. Whereas copy number gains peaking at 12q13.3 (STAT2, STAT6) (OR=0.38, p=0.0215) (Fig. S7E) and losses peaking at 7q34 (BRCA, EZH2, SMO) (OR=0.33, p=0.0285) (Fig. S7F) and 10q26.3 (MGMT) (OR=0.28, p=0.0148) (Fig. S7G) were associated with superior response. Both, BRCA loss^57^ and MGMT loss^58^ might predispose to increased sensitivity to DNA-damaging agents which were used as part of the patient’s treatment.

We did not observe a relationship between baseline cfDNA concentration in plasma and treatment response assessed by PET (Deauville 1-3 vs. Deauville 4-5: 13.4 vs. 18.3 ng/ml, p=0.5470) (Fig S7H).

Mutational burden (MB) was highly heterogeneous (Fig. S8A-B) and did not correlated with cfDNA concentration in plasma excluding a technical bias where high ctDNA amounts allow for detection of more variants (Fig. S8C). B2M mutations (10.0 vs. 5.9 mutations/Mb, p=0.0002) (Fig. S8D), HLA locus losses (8.6 vs. 5.9 mutations/Mb, p=0.0103) (Fig. S8E) and 9p24.1 gains (8.1 vs. 5.9 mutations/Mb, p=0.0343) (Fig. S8F) were associated with higher mutational burden, suggesting that high MB HL selected for these immune escape variants during development. In line with this, any mutation in MHC class I antigen presentation genes was associated with higher MB (9.4 vs. 4.1 mutations/Mb, p<0.0001) (Fig. S8G. These results suggest that a subset of HL exists that has a higher mutational burden and therefore immune escape variants had a selective advantage in its evolution and became dominant.

### Clonal structure of Hodgkin lymphoma

ctDNA sequencing can be used to deconvolute the spatial and temporal clonal structure of cancer. We used PyClone^59^ to differentiate between main clone mutations likely present in all HL cells and subclonal mutations likely only present in a fraction. Some OGs and TSGs harbored mainly main clone mutations (e.g. GNA13, XPO1, NFKBIE, IKBKB, CSF2RB, B2M), while some mainly subclonal mutations (e.g. PRBM1, NOTCH2, CHD2, BCR) (Fig. 5A). Some important HL OGs and TSGs (STAT6, SOCS1, TNFAIP3) harbored a mixture of main clone and subclonal mutations (Fig. 5A). Clonal structure allows inferring timing of mutations with main clone mutations likely occurring earlier in oncogenesis. Thus, the diversity in observed clonal structure suggests that mutational processes in HL oncogenesis are likely ongoing without a clear temporal order of genes important in early versus late oncogenesis.

**Figure 5:**
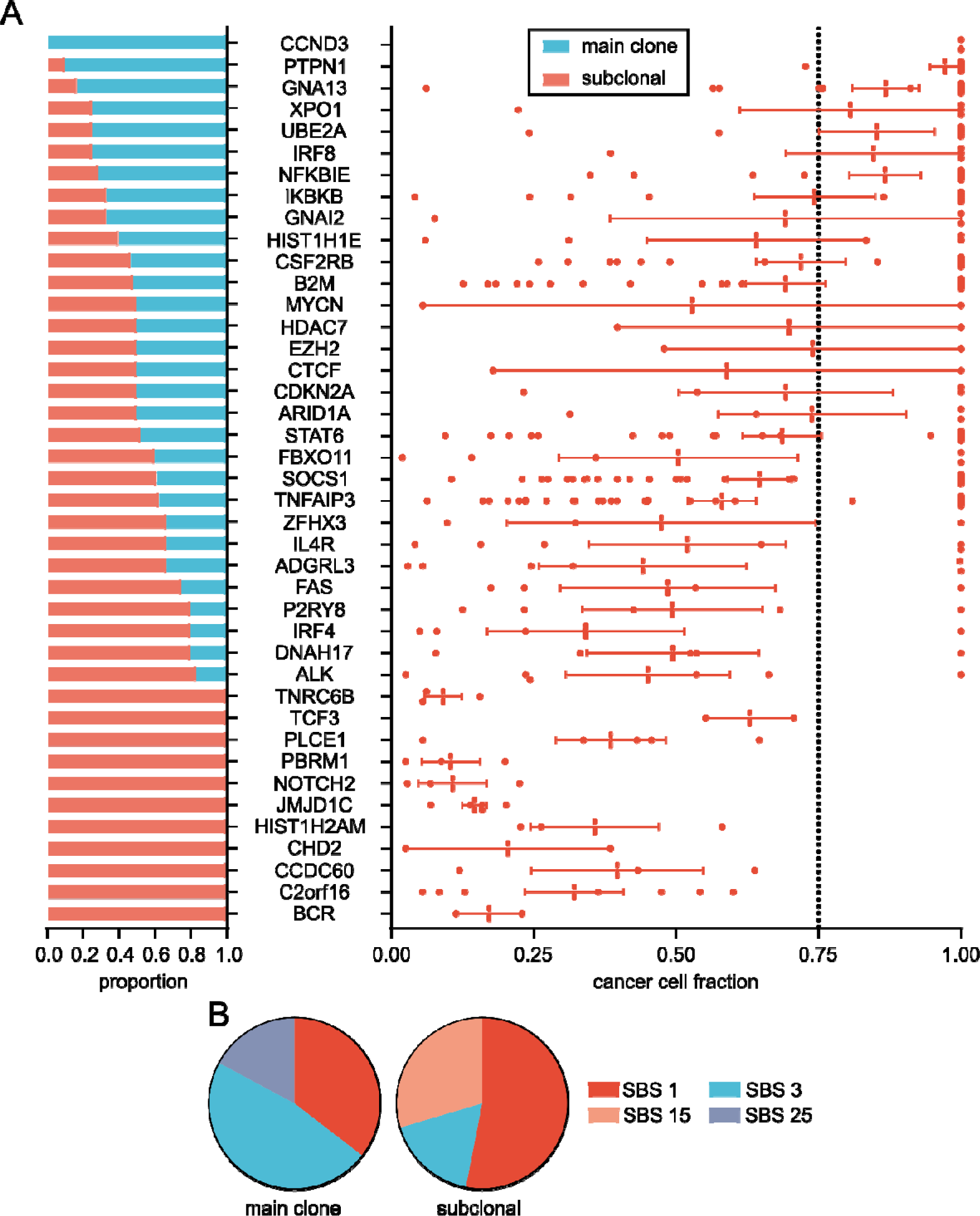
Clonal structure of Hodgkin lymphoma. (A), clonal structure is shown for all oncogenes and tumor suppressor genes (n=42) including all patients where clonal structure could be derived (n=51). The left chart shows percentage of patients with mutations in the indicated gene where at least one of the mutations in the indicated gene is in the main clone (cancer cell fraction (CCF) > 0.75). The right chart shows detailed CCFs. Each dot represents the CCF in the indicated gene in one patient where clonal structure could be assessed. The mean CCF for each gene is also shown, (B) contribution of single base substitution signatures to mutations in genes mainly mutated in main clones or subclones. Error bars show s.e.m.

To disentangle the contribution of mutational processes to early versus late HL oncogenesis, we compared mutational signatures in subclonal versus main clone mutations. SBS3 (defective HR), contributed more to main clone mutations compared to subclonal ones, whereas SBS15 (defective MMR) contributed more to subclonal mutations (Fig. 5B). This suggests that defective homologous recombination plays a bigger role in early HL oncogenesis, whereas defective DNA mismatch repair contributes more to ongoing mutational processes after initial malignant transformation.

### Minimal residual disease

Highly sensitive methods for response assessment that are predictive of treatment outcome are lacking in HL. Therefore, we aimed to use our ctDNA sequencing platform to measure MRD in HL, by following individual mutational *fingerprints* over time^15–17^. First, we examined the relationship between MRD at different timepoints and interim PET-imaging after 2 cycles of chemo-, immune- or combined chemoimmunotherapy in 43 follow-up samples^15^. MRD trajectories were markedly different between interim PET-response groups (Fig. 6A). Of note, MRD trajectories separated very early on, with clear differences already observable after 1 week (Fig. 6B). PET- and MRD-response measured after 2 cycles correlated in general (Fig. 6C-D) but differed sufficiently to underscore the potential for synergistic response assessment by PET and MRD as illustrated by individual patient MRD trajectories (Fig. 6E). Dichotomizing MRD assessment at the limit of detection, positive MRD after 1 week differentiated with 100% accuracy between Deauville 1-3 and Deauville 4-5 patients (Fig. 6F). Similarly, negative MRD after 2 cycles was associated with negative interim PET at the same timepoint (Fig. 6G).

**Figure 6:**
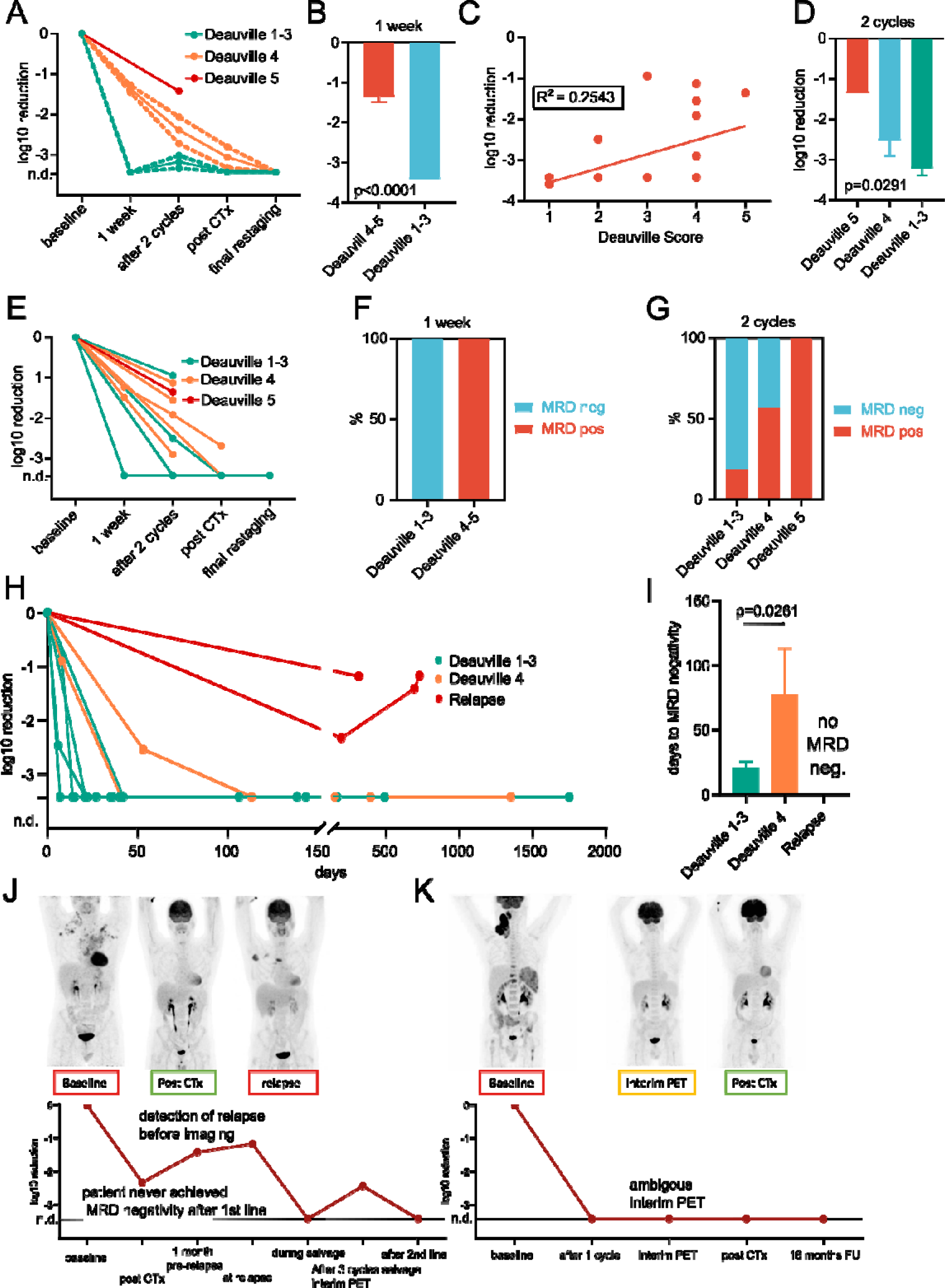
Detection of minimal residual disease. (A) mean reduction of minimal residual disease (MRD) during treatment depending on early interim PET-response (n=68 samples). Dotted lines indicate s.e.m., (B) difference in mean reduction of MRD 1 week after treatment initiation between interim PET-response groups (n=12 samples), (C) correlation between MRD after 2 cycles of treatment and time-matched interim PET-response (n=24 samples), (D) difference in mean reduction of MRD after 2 cycles of treatment between interim PET-response groups (n=48 samples), (E) single patient reduction of minimal residual disease (MRD) during treatment depending on early interim PET-response (n=68 samples). Each line represents MRD over time in one patient, (F) difference in MRD positivity or negativity 1 week after treatment between interim PET-response groups (n=12 samples), (G) difference in MRD positivity or negativity after 2 cycles of treatment between interim PET-response groups (n=48 samples), (H) single patient reduction of MRD in validation cohort. Each line represents MRD over time in one patient, (I) comparison of days to MRD negativity across patients in the validation cohort with either interim PET-negativity, interim PET-positivity, or relapse, (J-K) MRD and PET-imaging (PET images were also used for other studies that included the same patients) over time for two selected patients. The annotations below the PET-images correspond to the MRD timepoints named identically in the MRD chart. The color of the frame of the annotation indicates PET-positivity (red), ambiguity (yellow) or negativity (green). The line indicates MRD not detected. Error bars show s.e.m. T-test (B and I) and one-way ANOVA (D) were used. n.d.: MRD not detected.

To validate our findings in an independent cohort, we assessed MRD in 10 newly diagnosed HL patients with 54 samples collected and processed independently in another institution (Cancer Center Amsterdam). Sampling density was higher in the validation cohort allowing for more detailed time-course analysis of MRD. Interim-PET (2 cycles of chemotherapy) negative (Deauville 1-3) patients had quicker resolution of MRD compared to interim PET-positive (Deauville 4) patients (Fig. 6H-I). 2 patients experienced relapse and never achieved MRD-negativity with one patient each being PET-negative and positive after first-line treatment (Fig. 6H-I).

To showcase MRD monitoring with our assay, we highlight two patients. The first patient (Fig. 6J) is a 39-year-old male with advanced-stage HL treated with 6 cycles BEACOPPesc. At the end of treatment, PET showed a complete response, while MRD assessment was still positive. One and a half years after treatment, the patient relapsed. Both at relapse and, importantly, already 1 month prior to relapse, MRD assessment was positive. Compared to the MRD measurement directly after first-line treatment, the MRD level increased approximately 15-fold, even before imaging detected relapse. During second-line treatment (DHAP and autologous stem cell transplantation), the first MRD assessment was negative. At the end of salvage therapy, the patient was interim PET negative, however slightly MRD positive. After autologous stem cell transplantation, the patient remained in complete response and MRD assessment turned negative. This case highlights the possibility of our MRD assay to predict relapse after first-line treatment, detect it before imaging and aid in guiding treatment during second-line treatment. The second patient (Fig. 6K) is a 52-year-old male with advanced-stage HL treated with 6 cycles of BreECADD. MRD assessment was negative after 1 cycle and remained negative throughout treatment. PET based interim analyses, showed a discordance between local center assessment (complete metabolic response) and central image review (partial metabolic response), illustrating that MRD assessment may function as an auxiliary tool in more complex cases.

## Discussion

Here, we present the largest genomic landscape of HL to date and introduce our targeted ctDNA sequencing platform that allows for accurate genotyping using pre-treatment samples and highly sensitive MRD detection in samples collected during and after treatment. While applied to HL, this study serves as a blueprint for other cancers with similar therapeutic challenges.

The low tumor DNA content in bulk HL tissue due to the scarcity of HRS cells in primary lymph node biopsies^60^ has resulted in comparatively few genomic characterization studies in HL. Available knowledge is obtained from small studies with limited clinical data which relied on cumbersome sorting methods to enrich for HL-derived DNA. The three major studies in the field included only 10, 23 and 34 patients respectively^12, 33^. Reichel et al.^12^ and Wienand et al.^14^ used flow cytometry sorting to separate malignant from non-malignant cells in primary lymph node biopsies. Tiacci et al.^33^ used laser microdissection to achieve the same. Recently, two studies introduced ctDNA sequencing in HL as a tool to genotype adult or pediatric HL^22, 23^. Compared to both studies, our approach has several advantages. First, our target region is approximately 10x larger than the one used in both studies while at the same time, sequencing is performed at comparable depth. Second, our optimized bioinformatics process for error reduction and usage of UMIs results in highly reduced error rates, facilitating more accurate genotyping and orders of magnitude more sensitive MRD detection. Third, our bioinformatics platform allows for comprehensive genotyping including copy number variations and, if within the target region, structural variants. These advantages result in a higher proportion of patients with variants and a higher number of variants detected per patient. Therefore, we are convinced that our platform has the potential to become the state-of-the-art ctDNA pipeline in HL and in addition has wide applicability beyond HL only requiring modifications of the target region. For example, it has recently been shown that improved sequencing protocols and bioinformatics can facilitate highly accurate genotyping in lung cancer^25^ or diffuse-large B-cell lymphoma^26^. Importantly, our patient samples were collected in ancillary studies accompanying clinical trials across many treatment centers, showing that decentralized sample collection and centralized processing and analysis of liquid biopsies is feasible in hematological malignancies.

In our study, we made several novel findings with relevance to HL pathogenesis. First, we identify novel recurrent mutations in HL, such as Y641 EZH2 mutations and mutations in IRF4 and IRF8. Interestingly, Y641 EZH2 mutations have recently been shown to be able to modulate the germinal center niche towards lymphomagenesis, by reducing survival dependency of germinal center B-cells on T-cell help and driving expansion of germinal center centrocytes^61^. Second, we disentangle the contribution of different mutational processes in HL pathogenesis at high resolution, suggesting an important role of homologous recombination and offering first evidence that COSMIC SBS25 might indeed be a HL signature observable in patients. Third, we link, for the first time, HL genotypes to phenotypes and show that mutations in some genes, such as TP53 or NOTCH1, are associated with a high-risk phenotype, while other alterations, such as BRCA1 or MGMT loss are associated with a low-risk phenotype possibly mediated by increased sensitivity to commonly used cytotoxic drugs. Fourth, we identify a distinct genotype of large mediastinal mass HL with remarkable biological similarities to PMBL providing further evidence that an overlapping continuity between both entities exist while identifying large mediastinal mass HL for the first time as a HL subgroup more closely linked to PMBL than other classical HLs. Fifth, we classify HL as a disease with high heterogeneity of mutational burden and link high mutational burden HL with an immune escape genotype. It is possible that these patients will respond differently to immune checkpoint inhibition.

In a different use of our ctDNA sequencing platform, we show that it can be used for extremely sensitive MRD detection. It is especially worth mentioning that our MRD detection method can predict treatment response as early as one week after treatment initiation and predict relapse in PET-negative cases. Furthermore, MRD increases precede relapse detected by imaging. Other disease biomarkers, such as thymus and activation regulated chemokine (TARC)^62^ or exosomal miRNA^63^ have been evaluated in HL with some success, however patient numbers in these studies are low and preclude definitive conclusions. TARC has also been evaluated in conjunction with PET-CT imaging suggesting synergy^64^. ctDNA has been evaluated as a response biomarker in HL as well, for example in a study that looked at normalization of somatic copy number variation signatures in plasma^21^. However, it is likely that our approach is more sensitive as shown in the technical validation presented here. It is unlikely that other available methods for MRD detection in HL can detect a remaining plasma tumor burden of 0.00714% (less than 1 in 14,000 DNA molecules), although a head-to-head comparison is currently missing. In addition to synergy between MRD evaluation based on ctDNA sequencing and PET-CT, there might also be synergy between MRD evaluation based on ctDNA sequencing and protein-based biomarkers such as TARC in HL. The idea of combining protein-based and DNA-based biomarkers has been tested in other cancers with remarkable success^65^ and similar studies in HL are needed. Future enhancements of our assay could be the inclusion of *real* or in *silico* cfDNA size selection^66^, phased variant enrichment^67^ or duplex sequencing^68^.

In summary, we present the largest comprehensive genomic profiling study of HL to date utilizing our novel ctDNA sequencing platform and show that it can be used to accurately genotype HL at baseline and measure MRD with extremely high sensitivity. Our results pave the way for prospective clinical trials to evaluate prognostic genomic profiling by liquid biopsy and MRD-guided treatment in HL.

## Methods

### Samples and patients

324 samples from 121 patients were used in this study. 270 samples from 111 patients were used as the primary cohort in all analyses. 54 samples from 10 patients were used as a validation cohort in the MRD analyses only. Patients were de-identified. Samples for the primary cohort were collected within two German Hodgkin Study Group (GHSG) clinical trials: HD21 (n=44, ClinicalTrials.gov Identifier: NCT02661503) and NIVAHL (n=69, (ClinicalTrials.gov Identifier: NCT03004833). HD21 is a trial for newly diagnosed advanced stage HL patients. NIVAHL is a trial for newly diagnosed early-stage unfavorable HL patients^69^. Samples for the MRD validation cohort were routine care samples from Amsterdam University Medical Centers. Samples for the MRD validation cohort were collected within the BioLymph-study (2017-2019, VUmc METc registration number: 2017.008). The study is registered in the Dutch CCMO-register (toetsingonline.nl, NL60245.029.17). A part of this set of Amsterdam UMC samples (2014-2017), prior to the BioLymph study, has been collected through biobanking registered at the Biobank approval committee of VUmc, Amsterdam (2018.359).

### Ethical considerations

All patients provided written informed consent to allow the collection of peripheral blood for research purposes. All human subject research was performed in accordance with approved protocols by the local ethics committees and the Declaration of Helsinki.

### Statistical methods

Discrete variables were compared using Fisher’s exact test. Two continuous variables were compared using the student’s t-test if normality of variables could be assumed or the Mann-Whitney test if not. Multiple continuous variables were compared using one-way ANOVA with Welsh correction if normality of variables could be assumed or the Kruskal-Wallis test if not. Normality of variables was assessed heuristically examining the distribution of variable values. Statistical tests used are indicated when performed.

### Sample collection and processing

Blood samples were collected as part of an ancillary study to the main clinical trials. Samples were collected at predetermined time points within each trial. Samples were either collected in 6 or 9 ml EDTA tubes and processed within 2 hours, or in 10ml Streck cell-free DNA tubes (Streck, Omaha, NE) or 10ml PAXgene blood ccfDNA tubes (Qiagen, Hilden, Germany) and processed within 3 days. In case Streck cell-free DNA or PAXgene Blood ccfDNA tubes were used, they were first centrifuged at 1900g for 15 minutes at room temperature to separate the plasma. For further plasma purification, the plasma layer was pipetted into a clean 15ml LoBind tube (Eppendorf, Hamburg, Germany) and centrifuged at 1900 g for another 10 minutes. Plasma was then aliquoted into 2-5ml cryotubes and stored at −80°C. The cell pellet fraction remaining after the first centrifugation step was also aliquoted into 2-5ml cryotubes and stored along with plasma samples at −80°C for use as a germline control. In case EDTA tubes were used, the same protocol was followed, but the first centrifugation step was performed at 900g for 7 minutes at room temperature and the second centrifugation step was performed at 2500g for 10 minutes at room temperature.

### DNA extraction

To extract cell-free DNA from plasma samples, the QIAamp Circulating Nucleic-Acid Kit (Qiagen) was used according to manufacturer’s instructions but the Proteinase K incubation at 60°C was lengthened to 2 hours. All samples were extracted manually using a vacuum manifold (Qiagen). 2-4ml of plasma were used as input. To extract germline DNA from cell pellets, the Qiagen QIAamp DNA Blood Mini Kit (Qiagen) was used according to manufacturer’s instructions (spin protocol). 200µl of cell pellet was used as input. For DNA extraction from Formalin-Fixed Paraffin-Embedded (FFPE) lymph node samples 10 µm thick slides were cut. DNA extraction was done with the GeneRead DNA FFPE Kit (Qiagen) according to manufacturer’s instructions.

### DNA processing, quality control and sequencing

For target enrichment we used customized RNA baits designed with the SureSelect platform (Agilent, Santa Clara, CA). Our target panel has evolved throughout the study, so we used 3 different versions of RNA bait designs targeting 3.847 Mb (version 1), 2.981 Mb (version 2) and 2.997 Mb (version 3). Supplementary Table 11 shows which RNA bait design was used for which sample. Supplementary Table 12 shows the regions included in each design. For each sample, individual library preparations, hybridizations and captures were performed. For a first quantification of input DNA, the Qubit fluorometer (Invitrogen, Waltham, MA) was used. We used 25ng of DNA for library preparation where available but required a minimum amount of 15ng. For a more detailed quantification of input DNA considering the size distribution and as part of our quality control, the TapeStation 2200 System (Agilent) was used to quantify input DNA amounts in the size window of interest for cell-free DNA (50-700bp). We required a minimum of 25% of total input DNA to be in the correct size window. TapeStation DNA quantifications restricted to the size window of interest for cell-free DNA (50-700bp) were used for all further calculations requiring DNA amounts as input such as MRD quantification. Libraries were prepared using the SureSelectXT HS Automated Target Enrichment (Agilent) for Illumina paired-end multiplexed sequencing protocol including a unique molecular identifier (UMI) sequenced with an additional index read and the Agilent Bravo automated liquid handling platform (Agilent). After quality control using the 2200 TapeStation System and quantification using the Qubit System (Thermo Fisher, Waltham, MA), pools of libraries were generated. These pools were then sequenced on an Illumina NovaSeq6000 sequencing system (Illumina, San Diego, CA) using S1, S2 or S4 flow cells and 2 x 100 bp read length or an Illumina HiSeq4000 sequencing system (Illumina) with 2 x 75 bp read length.

### Basic data processing

Bcl2fastq2 software (v2.20, Illumina) was used to demultiplex bcl raw data. We used default settings to demultiplex the forward R1 and reverse R3 reads (--mask-short-adapter-reads 22 --minimum-trimmed-read-length 35 --barcode-mismatches 1 --adapter-stringency 0.9). Fastq files were trimmed with the bcl2fasq2 software. The UMI R2 reads were demultiplexed in a separate run using the following settings: --mask-short-adapter-reads 0 and --minimum-trimmed-read-length 0 without any adapter trimming. Raw reads were mapped to the human genome reference-build hg19 using the Burrows Wheeler Aligner (BWA) alignment algorithm with a base quality threshold of 15 for read trimming (parameter: -q 15)^70^. The resulting BAM files were further processed by a pipeline specifically designed for this project. Briefly, AddUMIsToBam (https://github.com/mbusby/AddUMIsToBam) was used to assign UMIs originating from every read to each aligned read in each BAM file. Fgbio’s (http://fulcrumgenomics.github.io/fgbio/) GroupReadsByUmi function was then used to group UMIs using the adjacency strategy with a maximum of 1 edit allowed. Next, Fgbio’s CallMolecularConsensusReads function was used to call a consensus read for each group of reads likely originating from the same molecule as identified by the same mapping and UMI with standard settings and accepting also one read of each unique input molecule to define that molecule’s sequence. Finally, reads are realigned using BWA mem using standard settings^70^. MPILEUP files are generated using Samtools^71^ ignoring overlaps (parameter: -x), adjusting mapping quality for reads containing excessive mismatches (parameter: -C 50), considering only reads with a minimum mapping quality of 40 (parameter: -q 40) and considering only bases with a minimum base quality of 30 (parameter: -Q 30). The MPILEUP file was limited to the genomic region captured during library preparation (parameter: -l).

### Somatic single base substitution calling

To improve sensitivity and specificity, somatic single base substitution calling was aided by comparative error suppression (CES). For this procedure, cell-free DNA from a cohort of 10 healthy donor patients was prepared and sequenced as described above. Post UMI-processing consensus reads aligned to hg19 by BWA mem^70^ were used as input. Using customized scripts, all positions with a non-reference (hg19) allele frequency > 5%, likely representing germline single nucleotide polymorphisms, were set to a non-reference allele frequency of 0 in the reference cohort. Next, TNER^27^ was used to compile a database of position-specific average background error rates for each position in the target region using the healthy donor controls as input. This database was used as input to TNER^27^ to polish each cell-free DNA MPILEUP generated from post UMI-processing consensus reads aligned to hg19 by BWA mem^70^ as described above with the polish strength parameter set to 0.05 (parameter: input.alpha = 0.05). Briefly, TNER^27^ uses tri-nucleotide error rates in the healthy reference cohort to suppress likely sequencing errors, thus reducing tri-nucleotide error rate profiles to suppress stochastic sequencing errors. Additionally, stereotypical sequencing errors at positions with high error rates were suppressed using customized scripts. To this end, each potential variant position was compared against the position-specific average background error rate profile generated above. If the average background error at a specific position was >0.5%, this position was blacklisted for variant calling as a position with a high rate of sequencing errors.

Next, this error-corrected dataset generated for each cell-free DNA sample was used as input for a customized variant calling pipeline. A variant was called at a position, when the following criteria were met: the minimum sequencing depth at a position was 50, at least 11 reads are supporting the variant, the variant has support on both the plus and the minus strand, the relative variant allele frequency of the variant is at least 0.1%, the variant has a relative variant allele frequency in the germline sample of less than 1%, the relative allele frequency in the cell-free DNA sample is at least 3 times larger than in the germline sample, a Fisher’s exact test on a 2 x 2 contingency table including the variant versus reference and the cell-free DNA versus the germline reference sample has a p-value < 0.05 and finally a one-tailed binominal test with the alternative hypothesis that the observed variant allele frequency is higher than the background error rate at that position has a p-value < 0.05. As an additional filter, we checked each variant for its frequency in ExAC^72^. Variants with a frequency above 0.01 in ExAC version 0.3^72^ were excluded. Furthermore, we excluded single base substitutions that were in close proximity (<6 base pairs) to an identified InDel. Variants were annotated and classified using ANNOVAR and databases ExAC version 0.3 (parameter: protocol -exac03) dbNFSP version 3.0a (parameter: -protocol dbnsfp30a) and dbSNP version 147 (parameter: -protocol avsnp147)^73^.

### Somatic small InDel calling

We used Varscan2^74^ to call small somatic insertions and deletions (InDels) and customized scripts for filtering identified variants. We used MPILEUP files generated as described above from post UMI-processing consensus reads aligned to hg19 by BWA mem^70^ as input. First, a comprehensive, unfiltered list of all potential InDels was generated using Varscan2^74^ with the following settings: minimum coverage of cell-free DNA sample file 1 (parameter: --min-coverage), minimum coverage of corresponding germline file 1 (parameter: --min-coverage-normal 1), minimum relative allele frequency 10^-9^ (parameter: --min-var-freq), minimum frequency to call a variant homozygous 0.75 (parameter: --min-freq-for-hom), minimum somatic p-value 0.99 (parameter: --somatic-p-value) and strand filter off (parameter: -- strand-filter 0). Of note, the parameters were deliberately chosen to be very loose as more stringent filtering was supposed to take place in the next step.

Using customized scripts, we filtered each raw InDel call file so only calls fulfilling the following criteria survived filtering: relative allele frequency of the matched germline < 0.5%, somatic p-value of call <0.01, absolute allele frequency of the matched germline < 4, not called as loss of heterozygosity by VarScan2^74^, at least 11 reads supporting the variant, at least one read on the plus strand and one read on the minus strand supporting the variant and minimum sequencing depth of 50 at the variant location in the cell-free DNA sample. As an additional filter, we checked each variant for its frequency in ExAC^72^. Variants with a frequency above 0.01 in ExAC version 0.3^72^ were excluded. Variants were annotated and classified using ANNOVAR and databases ExAC version 0.3 (parameter: protocol -exac03) dbNFSP version 3.0a (parameter: -protocol dbnsfp30a) and dbSNP version 147 (parameter: -protocol avsnp147)^73^.

### Somatic copy number calling

A pipeline was developed to identify somatic copy number changes based on CNVKit^46^. First, cell-free DNA from a cohort of 10 healthy donor patients was prepared and sequenced as described above. This healthy control cohort was used as a reference set to define an average technical background, against which somatic copy number changes of a specific sample were measured. The target region input for CNVKit^46^ was defined as the region that was covered by the RNA baits for the respective panel design. CNVKit’s batch function^46^ was then run for each cell-free DNA sample, using post UMI-processing consensus reads aligned to hg19 by BWA mem^70^ as input. Access data provided by CNVKit^46^ for hg19 and the drop low coverage (parameter: --drop-low-coverage) option were used in all runs. Standard parameters were used for all other settings. Segmented copy number calls were generated and an amplitude threshold of 0.02 or -0.02 was used to define a segment as a somatic copy number gain or loss, respectively. We did deliberately not try to infer absolute somatic copy number changes, because (I) cell-free DNA is not purely tumor-derived and, sometimes heavily, diluted by background germline DNA reducing sensitivity, and (II) cell-free DNA-derived somatic copy number changes represent an average of individual tumor cells somatic copy number states in the body which are, especially in the case of HL, known to be very variable between individual tumor cells^75, 76^.

To derive arm-level and focal recurrent and thus potentially biologically relevant somatic copy number changes, GISTIC 2.0.^47^ was used. Segmented copy number calls were used as input and GISTIC 2.0. was run including the whole sample cohort with the following settings: cytoband and gene location information was taken from hg19, gain threshold 0.02 (parameter: -ta), loss threshold 0.02 (parameter: -td), calculate significance by gene (parameter: -genegistic 1), including broad-level analysis (parameter: -broad 1), threshold to distinguish broad from focal events 0.98 (parameter: -brlen), confidence level 0.75 (parameter: -conf), q-value threshold 0.25 (parameter: -qvt), performing arm peel off (parameter: -armpeel 1) and collapse method extreme (parameter: -gcm). Standard parameters were used for all other settings. Broad arm-level plots, gain and loss q-plots and copy number heatmap plots were generated from resulting output using GISTIC’s^47^ built-in plotting functions.

### Additional variant filtering

For some analyses, mutations were only included if they occurred in the coding region of the genome. This is indicated in the respective analysis.

To identify recurrently mutated genes which are likely driver genes^32^ and thus relevant for HL pathogenesis either as tumor suppressor genes or as oncogenes, we applied a slightly modified version of the Vogelstein rule^32^, which is a deliberate choice for a rather conservative approach tending to underestimate cancer driver genes^32^. Furthermore, the 20/20 rule originally described by Vogelstein et al.^32^ and derived, ratiometric methods such as the 20/20+ algorithm have been shown to highly reliably predict functionally validated, known cancer driver genes and have a high stability across datasets^77^. We used customized scripts to identify a gene as a tumor suppressor gene, if the ratio of the combined number of splice site and nonsense mutations as well as frameshift insertions and deletions to all coding mutations was ≥ 0.25. A gene was identified as an oncogene if it was not a tumor suppressor gene and the ratio of mutations leading to an amino acid change that is shared with at least one other mutation leading to an amino acid change to all mutations leading to an amino acid change was ≥ 0.20. Evaluation of genes as potential tumor suppressor genes or oncogenes was performed for all genes with at least 5 mutations in the cohort.

### Technical and clinical validation and Quality control

For technical validation we spiked well-characterized reference DNA (NA12878)^28, 29^ fragmented to approximately cfDNA size into healthy donor cfDNA at proportions of 10%, 1%, 0.1%, 0.05%, 0.025%, 0.01% and 0%. Based on an allele frequency of 50% for heterozygous variants, these spike-in samples represented technical cfDNA validation samples with an expected mutated variant allele frequency of 5%, 0.5%, 0.05%, 0.025%, 0.0125% and 0.005%, respectively. Spike-ins of 5%, 0.5% and 0% were used to technically validate variant calling, whereas spike-ins of 0.1%, 0.05%, 0.025%, 0.01% and 0% were used to technically validate minimal residual disease detection. All spike-in samples were sequenced and bioinformatically processed as described above. Processing was identical to actual patient samples. For selected analyses UMI-based error suppression, CES or both were omitted. To calculate error profiles of sequencing with or without one or both error suppression methods, we used the 0% spike-in sample and randomly chose 1000 positions that were both not single nucleotide polymorphisms (SNPs) in NA12878 and the healthy donor sample – and should thus have zero non-reference bases detected at each position – and calculated each positions error rate as the combined variant allele frequency of all non-reference bases at the respective position. We modelled the error rate of different error suppression strategies as a decay process after sorting the error rates from high to low with Y denoting the modelled error rate, Y_0_ denoting the highest observed error rate, K the estimated decay rate and X the position in the high-to-low sorted error rate profile:

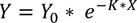

For technical validation of variant calling, we compiled a positive truth set of mutations (n=42) by identifying 22 cancer gene census genes^78^ that were (I) covered by our panel, (II) had at least one consensus variant identified in NA12878 and (III) were called in the 10% spike-in sample and thus in principle covered by our sequencing panel. Similarly, we compiled a negative truth set of non-mutated positions (n=1527) by randomly choosing one position in each genomic region covered by our panel of each of the 22 cancer gene census genes defined above that were both not single nucleotide polymorphisms (SNPs) in NA12878 and the healthy donor sample and including this position and both positions 3 bases up- and downstream, given they were also not single nucleotide polymorphisms (SNPs) in NA12878 and the healthy donor sample, into the negative truth set. The full positive and negative truth sets are available in Supplementary Table 10. We then performed variant calling in the 0.5% spike-in sample with or without UMI-based error suppression, CES or both and calculated sensitivity and specificity to detect variants with an allele frequency of 0.5% based on these truth sets.

For technical validation of minimal residual disease detection, we slightly modified the above-described positive truth set by limiting it to variants detected with an absolute variant allele frequency > 29 resulting in 40 variants. We calculated minimal residual disease as described above using this truth set as the technical validation baseline sample for the 0.1%, 0.05%, 0.025%, 0.01% and 0% spike-in samples with or without UMI-based error suppression, CES or both. To assess the contribution of UMI-based error suppression, CES or its combination to our assay’s sensitivity for minimal residual disease detection, we compared measured minimal residual disease in our technical validation spike-in samples with expected minimal residual disease based on expected mutated allele frequency in the spike-ins. We calculated bootstrap confidence intervals using R^79^ and the package boot^80^ based on 1000 bootstrap replicates. We calculated the limit of blank of our assay for minimal residual disease detection based on standard protocols for assay validation^30^ using the 0% spike-in sample as the blank sample and the 0.01% spike-in sample as a low-level sample. The limit of blank was defined as the mean blank minimal residual disease value + 1.645 x the 1000 replicate based bootstrap standard deviation of the blank sample’s mean minimal residual disease value. The limit of detection was defined as the mean blank minimal residual disease value + 1.645 x the 1000 replicate based bootstrap standard deviation of the low-level sample’s mean minimal residual disease value.

For clinical validation of variant calling, we subjected 8 corresponding Formalin-Fixed Paraffin-Embedded (FFPE) tumor biopsies to our sequencing and bioinformatics pipeline and compared called variants detected in the tumor biopsies with those detected in cfDNA.

For further clinical validation of the specificity of minimal residual disease detection, we tried to detect minimal residual disease in 10 healthy donor samples, which we used as minimal residual disease negative samples. We used variant call from 6 patients and subjected these 10 healthy donor samples to the standard minimal residual disease detection workflow using each patient’s variant call set, resulting in 60 independent experiments.

To calculate mean per gene coverage we calculated the mean coverage across all locations of single base substitutions in a given gene across the whole cohort. To calculate mean per patient coverage, we calculated the mean coverage across all locations of single base substitutions in a given patient.

### Clustering of mutations within 3-dimensional protein structures

Mutation3D^35^ was used to cluster variants within 3-dimensional protein structures in all identified oncogenes with at least 8 variants in our cohort as this was heuristically identified as the minimum number of variants needed to perform a meaningful analysis for the respective oncogene. For each analyzed oncogene, all amino acid sequence changes caused by identified variants where used as input to Mutation3D^35^, which was run in advanced mode. Settings were chosen to include all ModBase homology models with a ModPipe quality score (MPQS) ≥ 1.1, requiring at least three mutations and at least one unique amino acid change per cluster and allowing a maximum intracluster distance between mutations of 25Å. We considered an identified cluster sufficiently likely to be truly present if its p-value < 0.1.

### Mutual exclusivity and Co-occurrence of mutations analysis

To test if oncogenes or tumor suppressor genes co-occur or are mutually exclusive, we used DISCOVER^40^. We calculated the background matrix using all mutations and small InDels but limited our analysis to amino acid sequence altering mutations in oncogenes and tumor suppressor genes. We used a p-value of 0.05 as a threshold to detect mutual exclusivity or co-occurrence.

### Mutational burden

Mutational burden was calculated by dividing the number of mutations occurring within the target region for a given patient by the target region size.

### Mutational signatures

Mutational signatures were identified using R^79^ and deconstructSigs^42^. We deconstructed the mutations in each patient and for each gene with >30 mutations in our cohort into mutational signatures with the COSMIC single base substitution signatures as reference signatures^41^. Trinucleotide context normalization was based on trinucleotide context observations in the coding exome (parameter: tri.counts.method = ‘exome’). We considered a mutational signature detected in a patient or gene if its associated weight was > 0.1. Mutational signatures detected in at least 10% of patients in our cohort were considered to be recurrent mutational signatures present in HL (COSMIC single base substitution signatures 1, 3, 6, 9, 15 and 25). For both patients and genes, the proportion of mutations not explained by any of these signatures was classified as other/not assigned.

### Pathway associations

We assigned KEGG^81^, GO^82, 83^ or REACTOME^84^ pathways to identified oncogenes and tumor suppressor genes using g:Profiler^85^.

### Detection of viral and bacterial associations

To detect viral and bacterial associations of HL by liquid biopsy, we used a previously developed pipeline^48^ built around KRAKEN^49^. In brief, we used post UMI-processing consensus reads aligned to hg19 by BWA mem^86^ from all baseline cfDNA and germline control samples as input to KRAKEN^49^ and generated output reports which we filtered with KRAKEN’s built-in confidence scoring function with a confidence score of 0.25 (parameter: -- confidence 0.25). We used the KRAKEN standard database including viral and bacterial reference genomes for our analysis. Next, we arranged and filtered the output reports in a way that (II) excluded all human-aligned reads, (II) included only species-level hits, (III) excluded all species with > 4 cumulative alignments in our healthy control cohort to account for misalignments and common sequencing contaminants and (IV) excluded all species that had > 2 germline samples with > 4 reads aligned to it. In general, a species was determined to be detected in a cfDNA sample, if > 4 reads aligned to it. However, for EBV (HHV4) we used 33 as a threshold, as low-level EBV DNA is commonly detected in blood of healthy humans in highly sensitive assays^87^. To further exclude likely false-positive taxa, we excluded common sequencing contaminants compiled within one of our previous studies^48^, taxa that are common sources of biotechnological products (e.g. enzymes), such as bacteria from the Thermaceae family and thus also more likely to be contaminants and finally taxa that do not colonize mammalian hosts (See Supplementary Table 10 for details).

### Analysis of clonal structure of mutations

PyClone^59^ was used to analyze clonal structure of mutations. All identified single base substitutions in a patient were used as input. Normal copy number was set to 2. As absolute copy number information was not available, minor copy number and major copy number were set to 0 and 2, respectively. Prior was set to total copy number (parameter: --prior total_copy_number). Minimum cluster size was set to 3 (parameter: --min_cluster_size 3). Burnin was set to 1000 (parameter: --burnin 1000). Default settings were used for all other parameters. As a cleaning step, starting from the cluster with the highest mean cellular prevalence, clusters were pooled with the next cluster in order of decreasing mean cellular prevalence until a cluster size of at least 3 mutations was reached. Subsequent analyses of clonal structure were only performed for patients with at least 2 identified clones. To derive the cancer cell fraction (CCF) of each mutation, the cellular prevalence of the respective mutation was divided by the mean cellular prevalence of the cluster with highest mean cellular prevalence after cleaning as described above. The CCF of mutations in the main clone was set to 1. For subsequent dichotomized analyses, a mutation with a CCF > 0.75 was considered to be a main clone mutation and all other mutations were considered subclonal.

To analyze the relationship between mutational signatures and main clone versus subclonal mutations, deconstructSigs^42^ was run separately on all genes in which > 70% of mutations were main clone mutations (main clone dataset) and all genes in which > 80% of mutations were subclonal (subclone dataset).

### Minimal residual disease assessment

Minimal residual disease assessment was performed using customized R scripts. We used only single base substitution variant calls from baseline samples excluding non-coding RNA regions as input. Follow up samples were processed as described above including UMI-based and CES. Our process scans each follow up dataset at each variant position in the baseline dataset and identifies the sequencing depth and count of mutated alleles including only counts of the variant base identified in the baseline sample at each such position in the follow up sample. Then, it calculates *preliminary MRD,* defined as the sum of the count of all variant alleles defined as above in the follow up sample divided by the combined sequencing depth at these positions in the follow up sample. Next, we included a dusting step based on the assumption that mutated alleles should distribute evenly across all variants in the baseline file and outliers have a higher likelihood of being false tumor-derived somatic variant calls, for example CHIP variants^88^ not captured in the sample’s germline control leading to higher MRD detected than is truly present. We populated a Poisson distribution for each variant position with the expected number of events defined as: *preliminary MRD * sequencing depth at variant position in follow up sample*. We then derived the Poisson-likelihood that the observed or a higher number of mutated alleles at this position occurs. If this likelihood was < 10^-5^, we assumed this variant position had a higher than tolerable for MRD detection likelihood of being a false (e.g. non-tumor-derived) call and omitted it. After this dusting process we calculated final MRD as the sum of the remaining count of all variant alleles defined as above in the follow up sample divided by the combined remaining sequencing depth at these positions after the dusting process in the follow up sample.

To evaluate response in patients, we calculated the log_10_ reduction in MRD from baseline for a sample. We corrected for changes in cfDNA concentration between samples analogous to a method outlined before^89^. The log_10_ reduction was calculated according to the following formula:

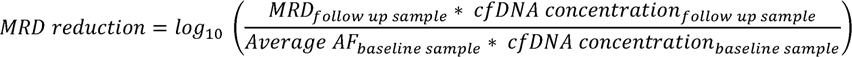

If MRD in a follow up sample was below the limit of blank, we denoted this as MRD not detected. For the purpose of statistical calculations within this manuscript we set MRD not detected to the midpoint between 0 and the MRD reduction of a fictitious *average* sample analogously to a previously described method^90^ with:

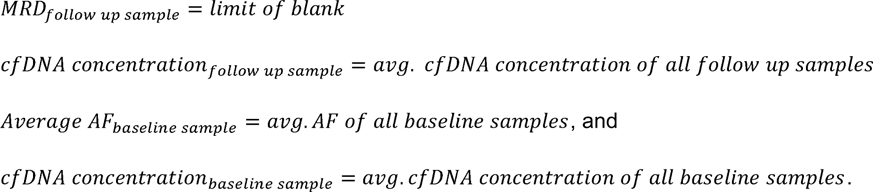

### Fluorescence in situ hybridization

FISH analyses were performed according to standard protocols. Briefly, bacterial artificial chromosome (BAC) clones were selected from the UCSC Genome Browser and ordered from Thermo Fisher. The BAC DNA was extracted from LB-Cultures according to standard protocols and labeled with Fluorescent Dye using the Nick Translation DNA Labeling System 2.0 (ENZO, New York, New York) according to manufacturer’s recommendations. The following BAC clones and labeled dUTPs (Fluorescent Dyes) were used: PD-L1/upstream: RP11-963L3 & RP11-12D24 (green 496 dUTP, ENZ-42831); PD-L2/downstream: RP11-207C16 & RP11-845C2 (orange 552 dUTP, ENZ-42842); centromere near control probe CTD-2024L1 & RP11-203L2 (aqua 431 dUTP, ENZ-42853).

To analyze genetic alterations in 9p24.1 we used a combination of CD30 IHC and FISH assay. We produced serial slides for CD30 IHC and FISH. For evaluation, we used the Bioview System (Abbott Molecular, Chicago, Illinois). We produced scans of the IHC slide and marked regions of interest (ROIs) containing large amounts of CD30-positive HRSC. Likewise, a scan of the FISH slide with the DAPI filter was recorded. Both of the scanned images (IHC and FISH) were matched at equivalent points and an overlay was produced. 50 tumor cells per case were analyzed in several of the previously selected ROIs.

## Data Availability

Primary sequencing data cannot be deposited publicly due to legal requirements and patient data protection. Upon entering a collaboration agreement and in consultation with the local ethics committee access to primary sequencing data might be granted. All secondary data derived from primary sequencing data is available within the article, supplementary information, or supplementary data files or available from the authors upon reasonable request.

## Code Availability

Custom scripts and code are available upon request.

## Acknowledgements

We would like to thank Elisabeth Kirst for excellent technical assistance, the staff at the trial sites of the included clinical studies for supporting this ancillary study and finally all patients, without whom this study would not have been possible.

---

**Supplementary Figure 1:**
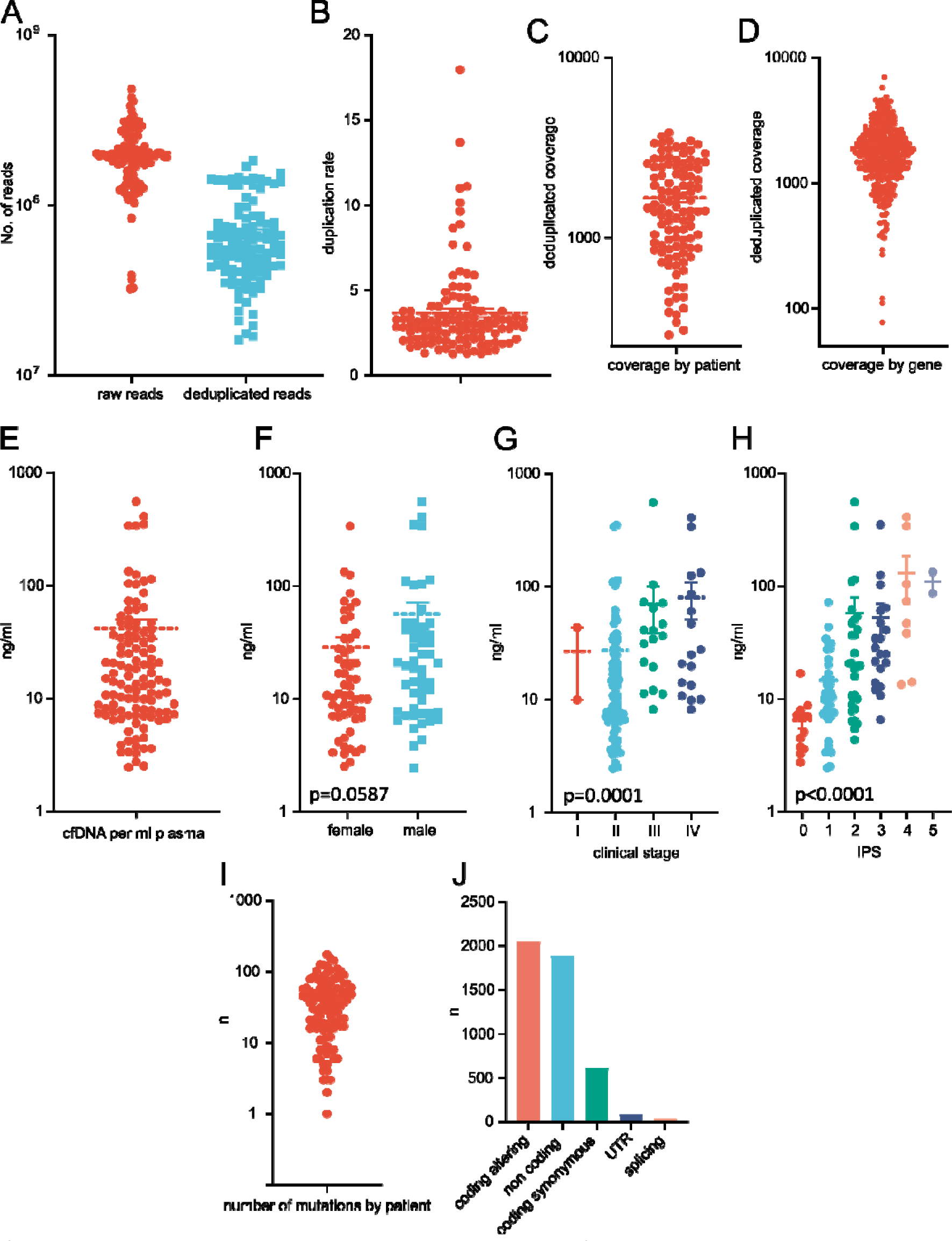
quality control measures, cfDNA and sequencing statistics. (A) distribution of raw read counts and uniquely mapped reads by patient (n=111), (B) duplication rate by patient (n=111), (C) distribution of deduplicated coverage by patient, (D) distribution of deduplicated coverage by gene (n=407), (E) distribution of cfDNA concentration in ng/ml of plasma (n=111), (F) distribution of cfDNA concentration by sex (n=111), (G) distribution of cfDNA concentration by clinical stage (n=111), (H) distribution of cfDNA amount concentration by international prognostic score (IPS) (n=111), (I) number of mutations detected by patient (n=109, 2 patients had no mutations detected), (J) absolute number of mutations of indicated categories detected in patient cohort (n=111). Error bars show s.e.m. Mann-Whitney test (F) and Kruskal-Wallis test (G-H) were used.

**Supplementary Figure 2:**
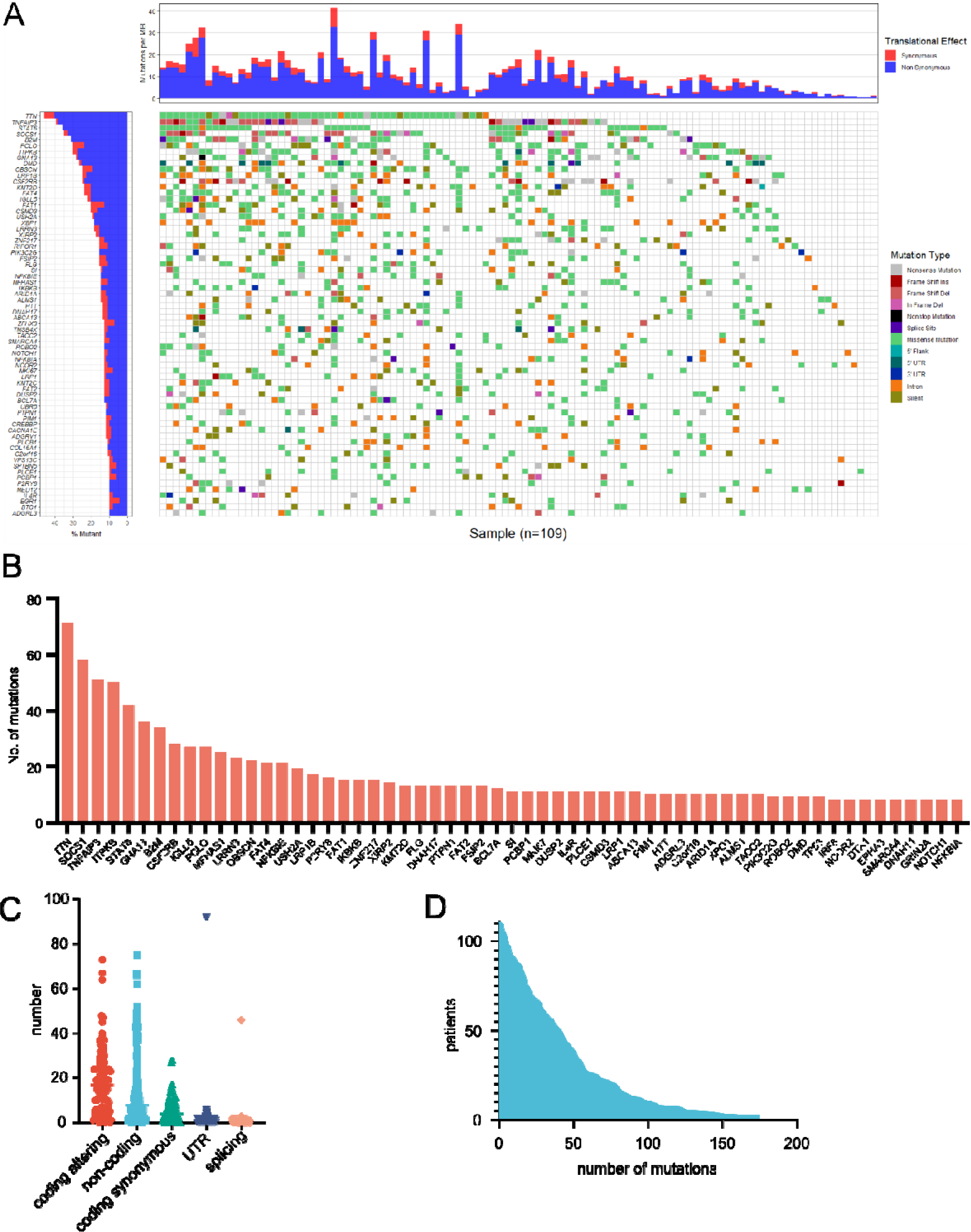
Mutational landscape of Hodgkin lymphoma (supplementing figure 2) (A) waterfall plot of most commonly mutated genes identified in the patient cohort (n=109, 2 patients without identified mutations not shown; miRNAs, ncRNAs and predominantly intronic mutated genes (BCL6, CIITA) - the intronic loci were intentionally targeted by our baits as common targets of AID – are excluded from plot). The chart on the left shows the genes and proportions of patients with mutations. The chart on top shows the mutational burden per patient. Color codes indicate the type of mutation in a patient and proportion of synonymous vs. non-synonymous mutations in genes (left chart) and patients (top chart), respectively. (B) most frequently mutated genes in patient cohort (n=111) excluding synonymous, intronic and intergenic mutations, (C) distribution of number of identified mutations of indicated type by patient in patient cohort (n=111), (D) cumulative plot of patients and number of mutations in any gene in patient cohort (n=111). Error bars show s.e.m.

**Supplementary Figure 3:**
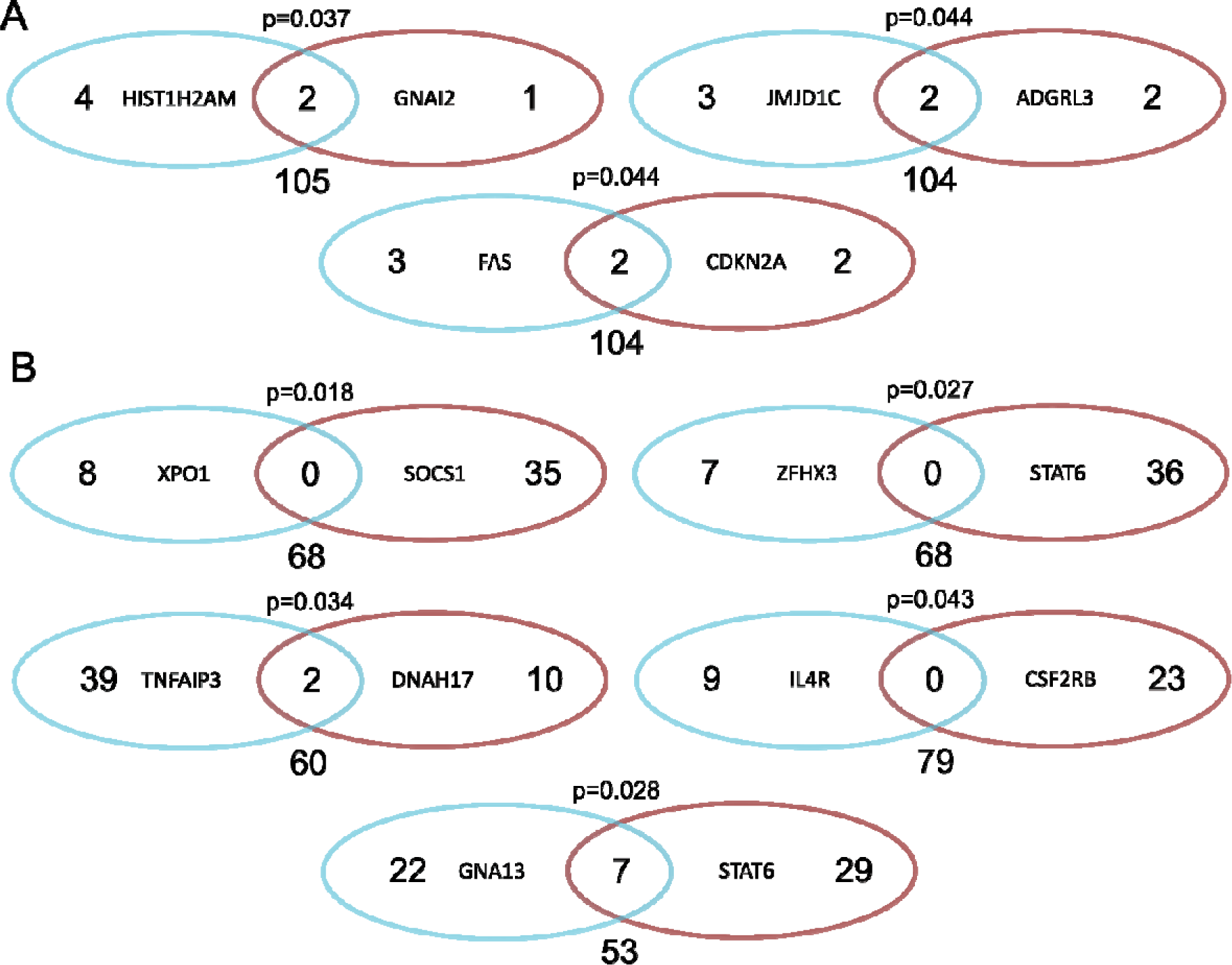
co-occurring and mutually exclusive mutations in Hodgkin lymphoma. (A) co-occurring mutations (n=111 patients), (B) mutually exclusive mutations (n=111 patients). The number in the overlap of the two oval shapes indicates the number of patients in which mutations in both indicated genes occur. The numbers in each of the oval shapes indicate the number of patients in which only a mutation in the indicated gene without a mutation in the other gene occurs. The number outside the two oval shapes indicate the number of patients without a mutation in any of the two indicated genes.

**Supplementary Figure 4:**
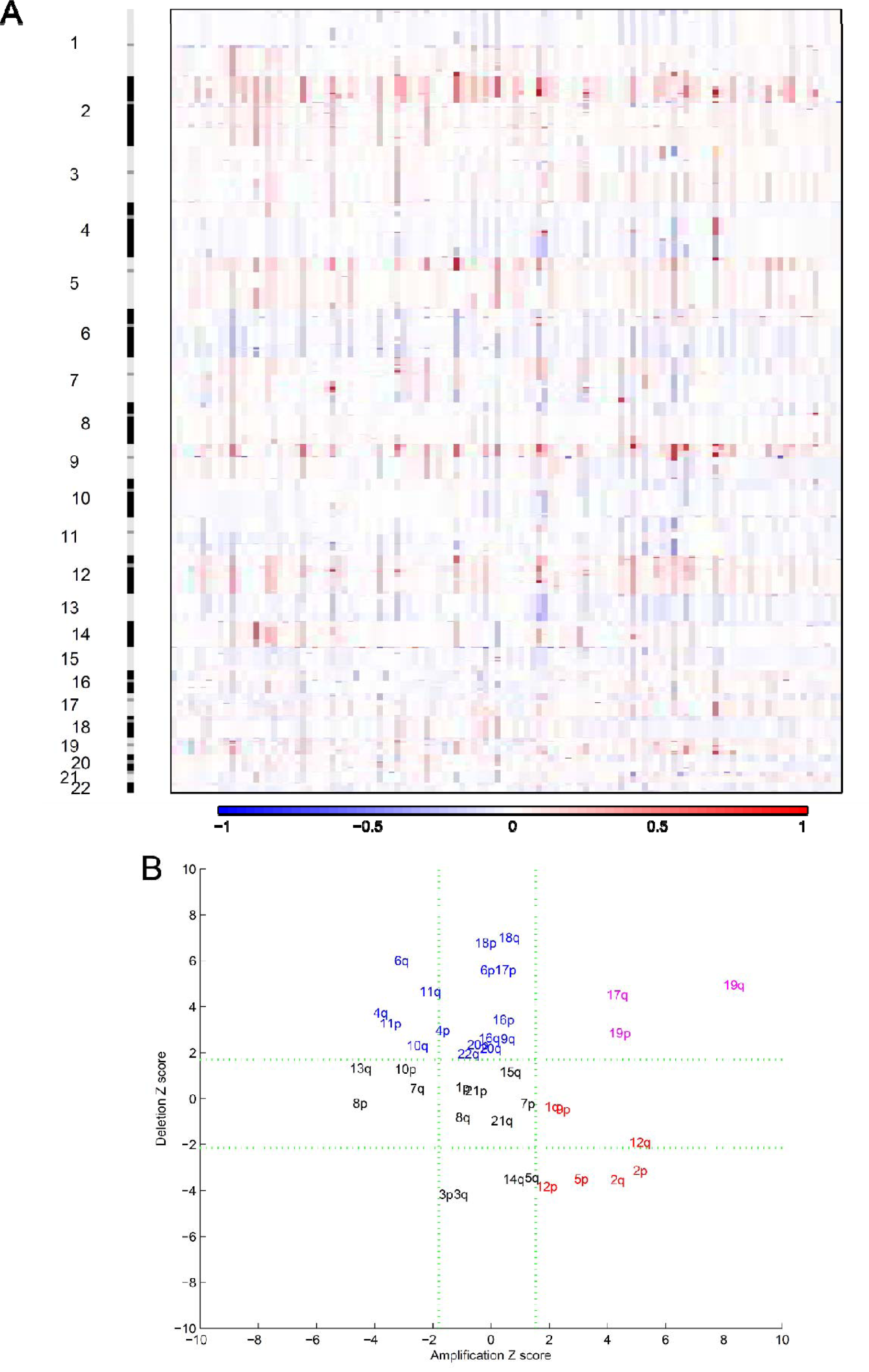
Copy number variations in Hodgkin lymphoma (supplementing figure 4) (A) plot of copy number profiles in each individual patient in patient cohort (n=111). The x-axis shows the chromosomal location, the y-axis shows the patients, (B) visualization of arm-wide copy number variations. Blue indicates recurrent arm-wide copy number losses, red indicates recurrent arm-wide copy number gains, pink indicates chromosomal arms that show both, recurrent copy number gains and losses.

**Supplementary Figure 5:**
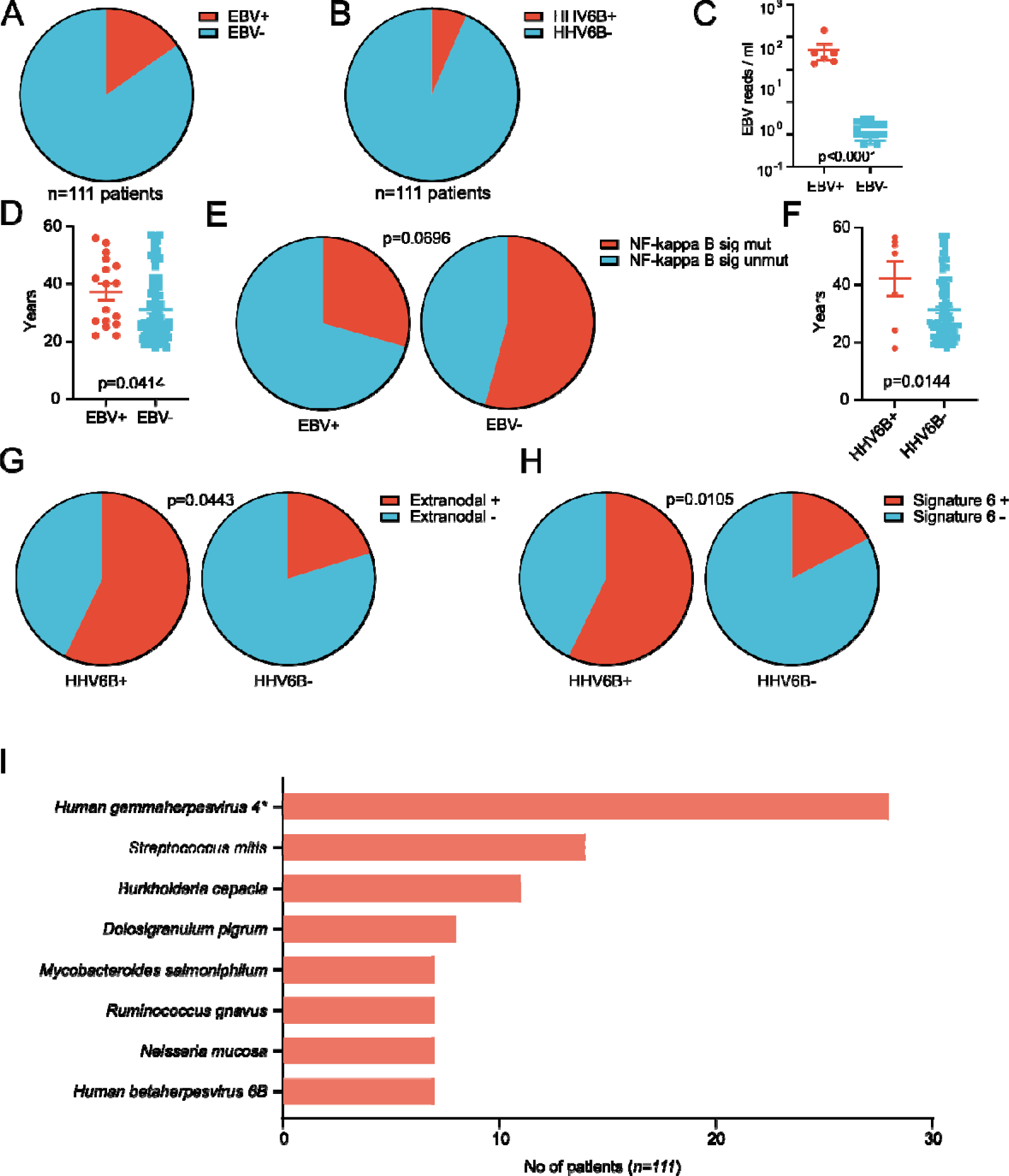
The plasma metagenome of Hodgkin lymphoma. (A) proportion of patients with detection of EBV above threshold (33 reads) in plasma (n=111 patients), (B) proportion of patients with detection of HHV6B above threshold (5 reads) in plasma (n=111 patients), (C) comparison of EBV reads / ml plasma between patients with and without detection of EBV in tumor cells of HL infiltrated lymph nodes by LMP1 immunohistochemistry (n=44), (D) comparison of patient age between patients with our without detection of EBV above threshold in plasma (n=111 patients), (E) comparison of proportion of patients with any mutation in the NF-kappa B signaling pathway in patients with or without detection of EBV above threshold in plasma (n=109 patients), (F) comparison of patient age between patients with our without detection of HHV6B above threshold in plasma (n=111 patients), (G) comparison of proportion of patients with extranodal disease in patients with or without detection of HHV6B above threshold in plasma (n=111 patients), (H) comparison of proportion of patients with significant detection of SBS 6 in patients with or without detection of HHV6B above threshold in plasma (n=109 patients), (I) most commonly detected viruses and bacteria in the plasma metagenome of the patient cohort (threshold 5 reads, n=111 patients). Error bars show s.e.m. T-test (C-D and F) and Fisher’s exact test (E and G-H) were used.

**Supplementary Figure 6:**
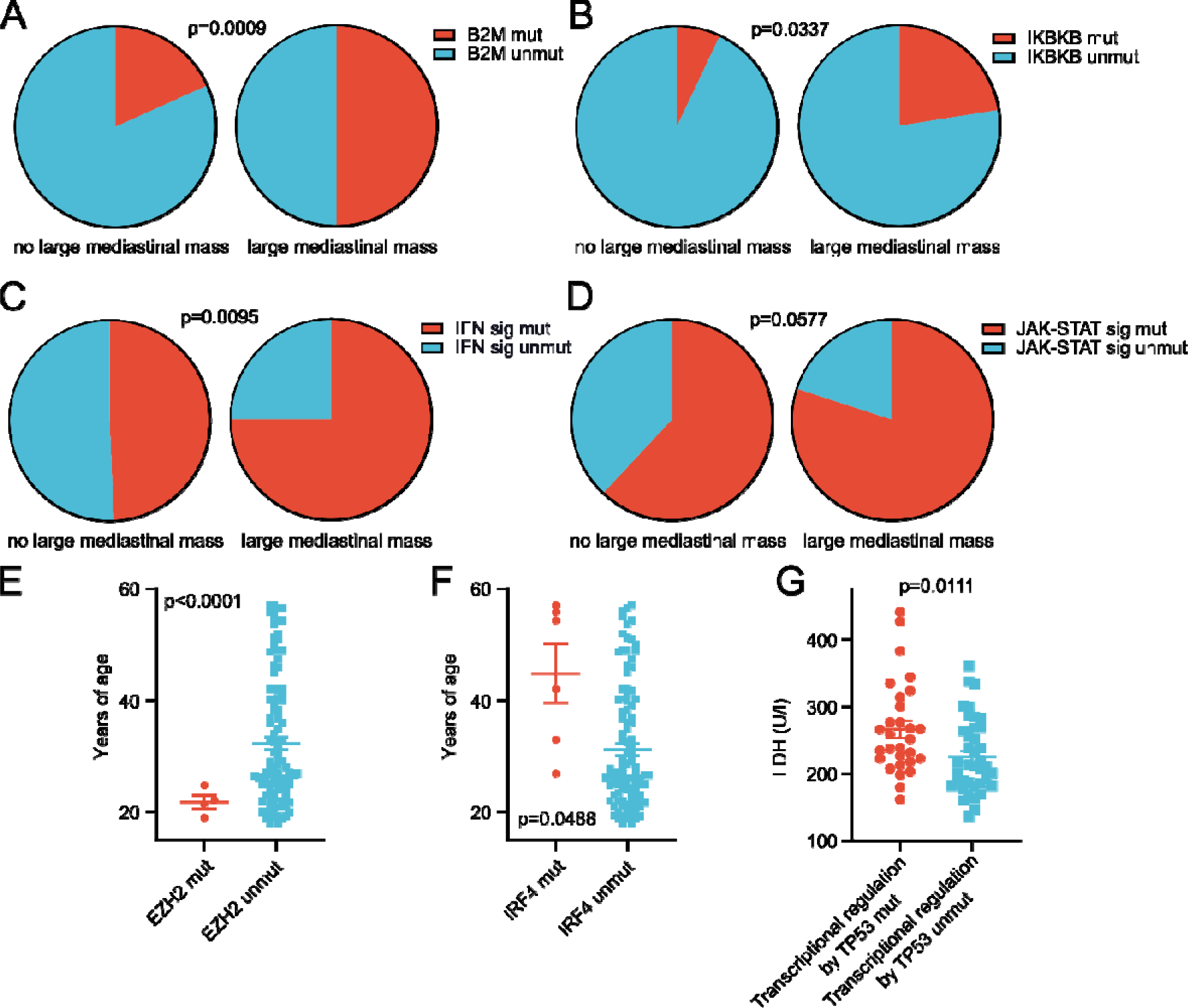
Genotype-phenotype associations in Hodgkin lymphoma. (A-B) comparison of proportion of patients with B2M (A) or IKBKB (B) mutations between patients with or without a large mediastinal mass (n=111 patients), (C-D) comparison of proportion of patients with any mutation in interferon signaling (C) or JAK-STAT signaling (D) between patients with or without a large mediastinal mass (n=111 patients), (E-F) comparison of patient age between patients with or without EZH2 (E) or IRF4 (F) mutations (n=111 patients), (G) comparison of lactate dehydrogenase (LDH) at diagnosis between patients with or without any mutation in genes involved in transcriptional regulation by TP53 (n=111 patients). Error bars show s.e.m. Fisher’s exact test (A-D) and t-tests (E-G) were used.

**Supplementary Figure 7:**
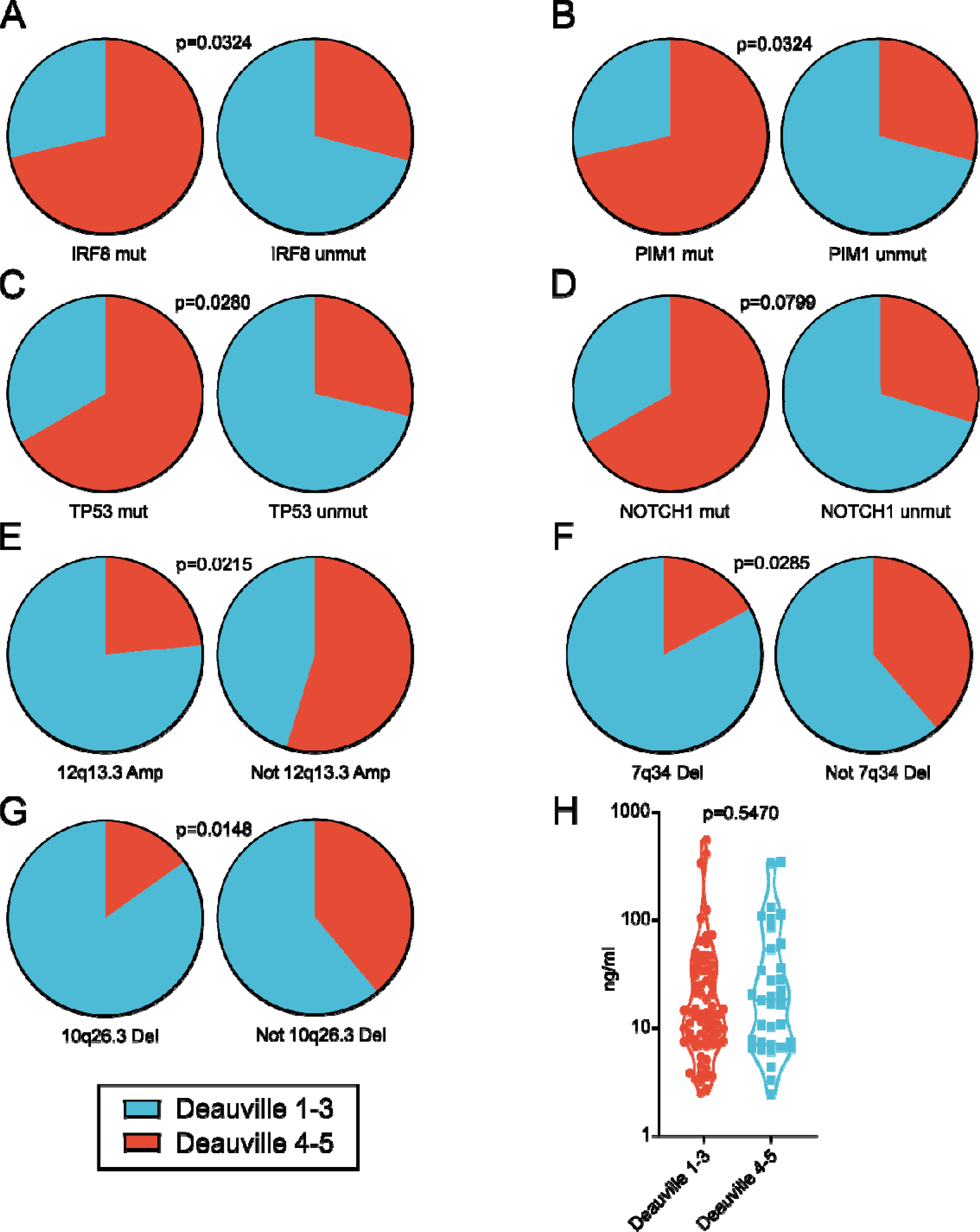
Association between genotypes and treatment response. (A-D) comparison of proportion of patients with IRF8 (A), PIM1 (B), TP53 (C) or NOTCH1 (D) mutations between patients with different early treatment response as indicated (n=111 patients), (E-G) comparison of proportion of patients with 12q13.3 gain (E), 7q34 loss (F) or 10q26.3 loss (G) between patients with different early treatment response as indicated (n=111 patients), (H) comparison of baseline cfDNA concentration between patients with different early treatment response as indicated (n=111 patients). Error bars show s.e.m. Fisher’s exact test (A-G) and t-test (H) were used.

**Supplementary Figure 8:**
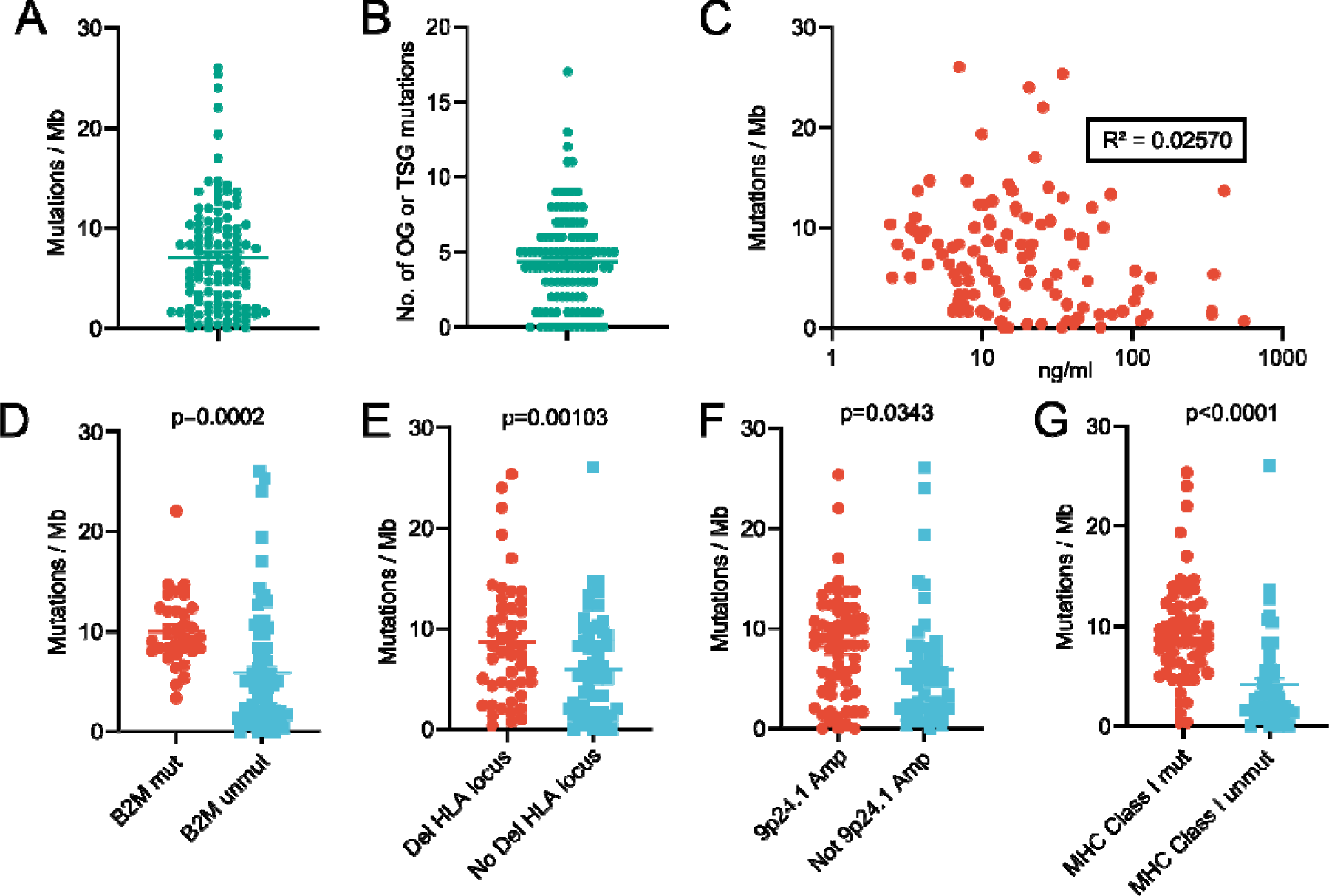
Mutational burden and associated genotypes. Mutational burden by patient (n=111), (B) absolute number of mutations in oncogenes or tumor suppressor genes by patient (n=111), (C) correlation between mutational burden and cfDNA concentration in baseline samples (n=111 patients), (D) comparison of mutational burden between patients with or without B2M mutations (n=111 patients), (E) comparison of mutational burden between patients with or without HLA locus loss (n=111 patients), (F) comparison of mutational burden between patients with or without 9p24.1 gain (n=111 patients), (G) comparison of mutational burden between patients with or without any mutation in genes involved in Class I MHC mediated antigen processing and presentation (n=111 patients). Error bars show s.e.m. T-tests (D-G) were used.

## Notes

Funding sources: SS was supported by a Mildred Scheel School of Oncology Aachen-Bonn-Cologne-Düsseldorf MD Research Stipend. Sequencing infrastructure was supported by DFG Research Infrastructure as part of the Next Generation Sequencing Competence Network [423957469]; NGS analyses were carried out at the production site WGGC Cologne. The research was partly funded by grant 70112502 from Deutsche Krebshilfe, grant EN 179/13-1 from the Deutsche Forschungsgemeinschaft (DFG) and a grant by the Frau-Weiskam und Christel Ruranski-Stiftung. It was also supported by the Hodgkin Lymphoma MRD consortium.

Conflict of Interest: PJB reports research grants from BeiGene, Bristol Myers Squibb, Merck Sharpe & Dohme, and Takeda; and personal fees and non-financial support from Bristol-Myers Squibb, Celgene and Takeda, all outside the submitted work. STS received travel grants from GSK. DMP reports being founder and CSO of Exbiome and an occasional advisor for Takeda. BvT reports personal fees and nonfinancial support from Bristol-Myers Squibb; personal fees from Amgen, Pfizer, Gilead Sciences, Pentixapharm, and Roche; grants, personal fees, and nonfinancial support from MSD and Takeda; and grants, personal fees, and nonfinancial support from Novartis. SB reports being founder, CEO and shareholder of Liqomics and personal fees and non-financial support from Bristol-Myers Squibb and Takeda outside the submitted work.

### Competing Interest Statement

PJB reports research grants from BeiGene, Bristol Myers Squibb, Merck Sharpe & Dohme, and Takeda; and personal fees and non-financial support from Bristol-Myers Squibb, Celgene and Takeda, all outside the submitted work. STS received travel grants from GSK. DMP reports being founder and CSO of Exbiome and an occasional advisor for Takeda. BvT reports personal fees and nonfinancial support from Bristol-Myers Squibb; personal fees from Amgen, Pfizer, Gilead Sciences, Pentixapharm, and Roche; grants, personal fees, and nonfinancial support from MSD and Takeda; and grants, personal fees, and nonfinancial support from Novartis. SB reports being founder, CEO and shareholder of Liqomics and personal fees and non-financial support from Bristol-Myers Squibb and Takeda outside the submitted work.

### Funding Statement

SS was supported by a Mildred Scheel School of Oncology Aachen-Bonn-Cologne-Duesseldorf MD Research Stipend. Sequencing infrastructure was supported by DFG Research Infrastructure as part of the Next Generation Sequencing Competence Network [423957469]; NGS analyses were carried out at the production site WGGC Cologne. The research was partly funded by grant 70112502 from Deutsche Krebshilfe, grant EN 179/13-1 from the Deutsche Forschungsgemeinschaft (DFG) and a grant by the Frau-Weiskam und Christel Ruranski-Stiftung. It was also supported by the Hodgkin Lymphoma MRD consortium.

### Author Declarations

All patients provided written informed consent to allow the collection of peripheral blood for research purposes. All human subject research was performed in accordance with approved protocols by the local ethics committees and the Declaration of Helsinki. The study was approved by the ethics committee of the University of Cologne (No. 16-008, 16-444 and 18-360) and the ethics committee of the VUmc Amsterdam (No. 2017.008 and 2018.359).

## References

1. Borchmann, S., von Tresckow, B. & Engert, A. Current developments in the treatment of early-stage classical Hodgkin lymphoma. Curr. Opin. Oncol. 28, 377–383 (2016).

2. Küppers, R., Engert, A. & Hansmann, M. L. Hodgkin lymphoma. Journal of Clinical Investigation 122, 3439–3447 (2012).

3. Kreissl, S. et al. Cancer-related fatigue in patients with and survivors of Hodgkin’s lymphoma: a longitudinal study of the German Hodgkin Study Group. Lancet Oncol. 17, 1453–1462 (2016).

4. Eichenauer, D. A. et al. Therapy-related acute myeloid leukemia and myelodysplastic syndromes in patients with hodgkin lymphoma: A report from the German Hodgkin Study Group. Blood 123, 1658–1664 (2014).

5. Borchmann, S. et al. Symptomatic osteonecrosis as a treatment complication in Hodgkin lymphoma: an analysis of the German Hodgkin Study Group (GHSG). Leukemia 33, 439 (2019).

6. Borchmann, S. et al. Thrombosis as a treatment complication in Hodgkin lymphoma patients: a comprehensive analysis of three prospective randomized German Hodgkin Study Group (GHSG) trials. Ann. Oncol. (2019).

7. Behringer, K. et al. Gonadal function and fertility in survivors after Hodgkin lymphoma treatment within the German Hodgkin study group HD13 to HD15 Trials. J. Clin. Oncol. 31, 231–239 (2013).

8. Borchmann, P., Eichenauer, D. a. & Engert, A. State of the art in the treatment of Hodgkin lymphoma. Nat. Rev. Clin. Oncol. 9, 450–459 (2012).

9. Hasenclever, D. & Diehl, V. A prognostic score for advanced Hodgkin’s disease. International Prognostic Factors Project on Advanced Hodgkin’s Disease. N. Engl. J. Med. 339, 1506–14 (1998).

10. Cirillo, M., Reinke, S., Klapper, W. & Borchmann, S. The translational science of hodgkin lymphoma. Br. J. Haematol. 184, 30–44 (2019).

11. Tiacci, E. et al. Pervasive mutations of JAK-STAT pathway genes in classical Hodgkin lymphoma. Blood 131, 2454–2465 (2018).

12. Reichel, J. et al. Flow sorting and exome sequencing reveal the oncogenome of primary Hodgkin and Reed-Sternberg cells. Blood 125, 1061–72 (2015).

13. Borchmann, S. & Engert, A. The genetics of Hodgkin lymphoma. Curr. Opin. Oncol. 1 (2017). doi:10.1097/CCO.0000000000000396

14. Wienand, K. et al. Genomic analyses of flow-sorted Hodgkin Reed-Sternberg cells reveal complementary mechanisms of immune evasion. Blood Adv. 3, 4065–4080 (2019).

15. Borchmann, P. et al. PET-guided treatment in patients with advanced-stage Hodgkin’s lymphoma (HD18): final results of an open-label, international, randomised phase 3 trial by the German Hodgkin Study Group. Lancet (London, England) 390, 2790–2802 (2017).

16. Stephens, D. M. et al. Five-year follow-up of SWOG S0816: Limitations and values of a PET-adapted approach with stage III/IV Hodgkin lymphoma. Blood 134, 1238–1246 (2019).

17. Gallamini, A. et al. Early chemotherapy intensification with escalated beacopp in patients with advanced-stage hodgkin lymphoma with a positive interim positron emission tomography/computed tomography scan after two abvd cycles: Long-term results of the GITIL/FIL HD 0607 trial. J. Clin. Oncol. 36, 454–462 (2018).

18. Döhner, H. et al. Diagnosis and management of AML in adults: 2017 ELN recommendations from an international expert panel. Blood 129, 424–447 (2017).

19. Bassan, R. et al. A systematic literature review and metaanalysis of minimal residual disease as a prognostic indicator in adult B-cell acute lymphoblastic leukemia. Haematologica 104, 2028–2039 (2019).

20. Newman, A. M. et al. An ultrasensitive method for quantitating circulating tumor DNA with broad patient coverage. Nat. Med. 20, 548–554 (2014).

21. Vandenberghe, P. et al. Non-invasive detection of genomic imbalances in Hodgkin/Reed-Sternberg cells in early and advanced stage Hodgkin’s lymphoma by sequencing of circulating cell-free DNA: a technical proof-of-principle study. Lancet Haematol. 2, e55–e65 (2015).

22. Spina, V. et al. Circulating tumor DNA reveals genetics, clonal evolution and residual disease in classical Hodgkin lymphoma. Blood blood-2017-11-812073 (2018). doi:10.1182/blood-2017-11-812073

23. Desch, A. K. et al. Genotyping circulating tumor DNA of pediatric Hodgkin lymphoma. Leukemia (2019). doi:10.1038/s41375-019-0541-6

24. Hohaus, S. et al. Cell-free circulating DNA in Hodgkin’s and non-Hodgkin’s lymphomas. Ann. Oncol. 20, 1408–13 (2009).

25. Newman, A. M. et al. Integrated digital error suppression for improved detection of circulating tumor DNA. Nat. Biotechnol. 34, 547–555 (2016).

26. Kurtz, D. M. et al. Circulating Tumor DNA Measurements As Early Outcome Predictors in Diffuse Large B-Cell Lymphoma. J. Clin. Oncol. 36, 2845–2853 (2018).

27. Deng, S. et al. TNER: A novel background error suppression method for mutation detection in circulating tumor DNA. BMC Bioinformatics 19, 387 (2018).

28. Zook, J. M. et al. An open resource for accurately benchmarking small variant and reference calls. Nat. Biotechnol. 37, 561–566 (2019).

29. Krusche, P. et al. Best practices for benchmarking germline small-variant calls in human genomes. Nat. Biotechnol. 37, 555–560 (2019).

30. Armbruster, D. A. & Pry, T. Limit of blank, limit of detection and limit of quantitation. Clin. Biochem. Rev. 29 **Suppl 1**, S49–52 (2008).

31. Lawrence, M. S. et al. Mutational heterogeneity in cancer and the search for new cancer-associated genes. Nature 499, 214–218 (2013).

32. Vogelstein, B. et al. Cancer genome landscapes. Science (80-.). 340, 1546–1558 (2013).

33. Tiacci, E. et al. Pervasive mutations of JAK-STAT pathway genes in classical Hodgkin lymphoma. Blood 131, 2454–2465 (2018).

34. Schmitz, R. et al. TNFAIP3 (A20) is a tumor suppressor gene in Hodgkin lymphoma and primary mediastinal B cell lymphoma. J. Exp. Med. 206, 981–989 (2009).

35. Meyer, M. J. et al. mutation3D: Cancer Gene Prediction Through Atomic Clustering of Coding Variants in the Structural Proteome. Hum. Mutat. 37, 447–456 (2016).

36. Cardinez, C. et al. Gain-of-function IKBKB mutation causes human combined immune deficiency. J. Exp. Med. 215, 2715–2724 (2018).

37. Cherian, M. A. et al. An activating mutation of interferon regulatory factor 4 (IRF4) in adult T-cell leukemia. J. Biol. Chem. 293, 6844–6858 (2018).

38. Baus, D. et al. STAT6 and STAT1 are essential antagonistic regulators of cell survival in classical Hodgkin lymphoma cell line. Leukemia 23, 1885–1893 (2009).

39. Taylor, J. et al. Altered nuclear export signal recognition as a driver of oncogenesis. Cancer Discov. 9, 1452–1467 (2019).

40. Canisius, S., Martens, J. W. M. & Wessels, L. F. A. A novel independence test for somatic alterations in cancer shows that biology drives mutual exclusivity but chance explains most co-occurrence. Genome Biol. 17, 261 (2016).

41. Alexandrov, L. B. et al. The repertoire of mutational signatures in human cancer. Nature 578, 94–101 (2020).

42. Rosenthal, R., McGranahan, N., Herrero, J., Taylor, B. S. & Swanton, C. deconstructSigs: Delineating mutational processes in single tumors distinguishes DNA repair deficiencies and patterns of carcinoma evolution. Genome Biol. 17, 31 (2016).

43. Petljak, M. et al. Characterizing Mutational Signatures in Human Cancer Cell Lines Reveals Episodic APOBEC Mutagenesis. Cell 176, 1282–1294.e20 (2019).

44. Duke, J. L. et al. Multiple Transcription Factor Binding Sites Predict AID Targeting in Non-Ig Genes. J. Immunol. 190, 3878–3888 (2013).

45. Schmitz, R. et al. *TNFAIP3* (A20) is a tumor suppressor gene in Hodgkin lymphoma and primary mediastinal B cell lymphoma. J. Exp. Med. 206, 981–989 (2009).

46. Talevich, E., Shain, A. H., Botton, T. & Bastian, B. C. CNVkit: Genome-Wide Copy Number Detection and Visualization from Targeted DNA Sequencing. PLOS Comput. Biol. 12, e1004873 (2016).

47. Mermel, C. H. et al. GISTIC2.0 facilitates sensitive and confident localization of the targets of focal somatic copy-number alteration in human cancers. Genome Biol. 12, R41 (2011).

48. Borchmann, S. An atlas of bacterial and viral associations in cancer. bioRxiv 773200 (2019).

49. Wood, D. E. & Salzberg, S. L. Kraken: ultrafast metagenomic sequence classification using exact alignments. Genome Biol. 15, R46 (2014).

50. Jarrett, R. F., Hjalgrim, H. & Murray, P. G. The Role of Viruses in the Genesis of Hodgkin Lymphoma. in Hodgkin Lymphoma: A Comprehensive Overview (eds. Engert, A. & Younes, A.) 25–45 (Springer International Publishing, 2020). doi:10.1007/978-3-030-32482-7_2

51. Jarrett, A. F., Armstrong, A. A. & Alexander, E. Review Epidemiology of EBV and Hodgkin’s lymphoma. Proceedings of the Third International Symposium on Hodgkin’s Lymphoma, September 19–23, 1995 - Köln, Germany 7, (Kluwer Academic Publishers, 1995).

52. Wei, W. et al. A20 and RBX1 Regulate Brentuximab Vedotin Sensitivity in Hodgkin Lymphoma Models. Clin. Cancer Res. 26, 4093–4106 (2020).

53. Han, X. Y., Kamana, M. & Rolston, K. V. I. Viridans streptococci isolated by culture from blood of cancer patients: Clinical and microbiologic analysis of 50 cases. J. Clin. Microbiol. 44, 160–165 (2006).

54. Thurner, L. et al. Lymphocyte predominant cells detect Moraxella catarrhalis-derived antigens in nodular lymphocyte-predominant Hodgkin lymphoma. Nat. Commun. 11, (2020).

55. Chapuy, B. et al. Genomic analyses of PMBL reveal new drivers and mechanisms of sensitivity to PD-1 blockade. Blood 134, 2369–2382 (2019).

56. Traverse-Glehen, A. et al. Mediastinal gray zone lymphoma: The missing link between classic Hodgkin’s lymphoma and mediastinal large B-cell lymphoma. Am. J. Surg. Pathol. 29, 1411–1421 (2005).

57. Lord, C. J. & Ashworth, A. BRCAness revisited. Nat. Rev. Cancer 16, 110–120 (2016).

58. Hegi, M. E. et al. MGMT Gene Silencing and Benefit from Temozolomide in Glioblastoma. N. Engl. J. Med. 352, 997–1003 (2005).

59. Roth, A. et al. PyClone: statistical inference of clonal population structure in cancer. Nat. Methods 11, 396–398 (2014).

60. Borchmann, S. & Engert, A. The genetics of Hodgkin lymphoma: an overview and clinical implications. Curr. Opin. Oncol. 29, 307–314 (2017).

61. Béguelin, W. et al. Mutant EZH2 Induces a Pre-malignant Lymphoma Niche by Reprogramming the Immune Response. Cancer Cell 37, 655–673.e11 (2020).

62. Sauer, M. et al. Baseline serum TARC levels predict therapy outcome in patients with Hodgkin lymphoma. Am. J. Hematol. 88, 113–5 (2013).

63. van Eijndhoven, M. A. J. et al. Plasma vesicle miRNAs for therapy response monitoring in Hodgkin lymphoma patients. JCI insight 1, e89631 (2016).

64. Plattel, W. J. et al. Interim thymus and activation regulated chemokine *versus* interim^18^ F fluorodeoxyglucose positron emission tomography in classical Hodgkin lymphoma response evaluation. Br. J. Haematol. 190, 40–44 (2020).

65. Cohen, J. D. et al. Detection and localization of surgically resectable cancers with a multi-analyte blood test. Science eaar3247 (2018). doi:10.1126/science.aar3247

66. Mouliere, F. et al. Selecting Short DNA Fragments In Plasma Improves Detection Of Circulating Tumour DNA. doi.org 134437 (2017). doi:10.1101/134437

67. Kurtz, D. M. et al. Phased Variant Enrichment for Enhanced Minimal Residual Disease Detection from Cell-Free DNA. Blood 134, 552–552 (2019).

68. Kennedy, S. R. et al. Detecting ultralow-frequency mutations by Duplex Sequencing. Nat. Protoc. 9, 2586–2606 (2014).

69. Bröckelmann, P. J. et al. Efficacy of Nivolumab and AVD in Early-Stage Unfavorable Classic Hodgkin Lymphoma: The Randomized Phase 2 German Hodgkin Study Group NIVAHL Trial. JAMA Oncol. 6, 872–880 (2020).

70. Li, H. & Durbin, R. Fast and accurate short read alignment with Burrows-Wheeler transform. Bioinformatics 25, 1754–1760 (2009).

71. Li, H. et al. The Sequence Alignment/Map format and SAMtools. Bioinformatics 25, 2078–2079 (2009).

72. Lek, M. et al. Analysis of protein-coding genetic variation in 60,706 humans. Nature 536, 285–291 (2016).

73. Sherry, S. T. et al. DbSNP: The NCBI database of genetic variation. Nucleic Acids Res. 29, 308–311 (2001).

74. Koboldt, D. C. et al. VarScan 2: Somatic mutation and copy number alteration discovery in cancer by exome sequencing. Genome Res. 22, 568–576 (2012).

75. Joos, S. et al. Hodgkin’s lymphoma cell lines are characterized by frequent aberrations on chromosomes 2p and 9p including *REL* and *JAK2*. Int. J. Cancer 103, 489–495 (2003).

76. Roemer, M. G. M. et al. PD-L1 and PD-L2 Genetic Alterations Define Classical Hodgkin Lymphoma and Predict Outcome. J. Clin. Oncol. 34, 2690–7 (2016).

77. Tokheim, C. J., Papadopoulos, N., Kinzler, K. W., Vogelstein, B. & Karchin, R. Evaluating the evaluation of cancer driver genes. Proc. Natl. Acad. Sci. U. S. A. 113, 14330–14335 (2016).

78. Sondka, Z. et al. The COSMIC Cancer Gene Census: describing genetic dysfunction across all human cancers. Nat. Rev. Cancer 18, 696–705 (2018).

79. Core Team, R. R: A Language and environment for statistical computing. R Foundation for Statistical Computing, Vienna, Austria. URL: https://www.R-project.org/. (2020).

80. Canty, A. & Ripley, B. boot: Bootstrap R (S-Plus) Functions. (2020).

81. Kanehisa, M., Sato, Y., Kawashima, M., Furumichi, M. & Tanabe, M. KEGG as a reference resource for gene and protein annotation. Nucleic Acids Res. 44, D457– D462 (2016).

82. Ashburner, M. et al. Gene ontology: tool for the unification of biology. The Gene Ontology Consortium. Nat. Genet. 25, 25–9 (2000).

83. The Gene Ontology Resource: 20 years and still GOing strong. Nucleic Acids Res. 47, D330–D338 (2019).

84. Fabregat, A. et al. Reactome pathway analysis: a high-performance in-memory approach. BMC Bioinformatics 18, 142 (2017).

85. Raudvere, U. et al. G:Profiler: A web server for functional enrichment analysis and conversions of gene lists (2019 update). Nucleic Acids Res. 47, W191–W198 (2019).

86. Li, H. & Durbin, R. Fast and accurate short read alignment with Burrows-Wheeler transform. Bioinformatics 25, 1754–1760 (2009).

87. Stevens, S. J. C. et al. Monitoring of Epstein-Barr virus DNA load in peripheral blood by quantitative competitive PCR. J. Clin. Microbiol. 37, 2852–2857 (1999).

88. Jaiswal, S. & Ebert, B. L. Clonal hematopoiesis in human aging and disease. Science 366, (2019).

89. Kurtz, D. M. et al. Circulating tumor DNA measurements as early outcome predictors in diffuse large B-cell lymphoma. J. Clin. Oncol. 36, 2845–2853 (2018).

90. Environmental Protection Agency. Guidance for data quality assessment: Practical methods for data analysis. (2000).

